# Substantia Nigra and Subthalamic Nucleus Deep Brain Stimulation Exert Opposing Effects on Novelty Recognition in Parkinson’s Disease

**DOI:** 10.64898/2026.06.17.26355856

**Authors:** Bingxin Li, Chenguan Jiang, Yan Liu, Zhou Yang, Anbang Yao, Xiaoyu Zhang, Yuye Liu, Hutao Xie, Barbara Hollunder, Bryan Strange, Anchao Yang, Fangang Meng, Jianguo Zhang, Changming Wang, Ningfei Li, Lin Shi

## Abstract

Episodic memory plays a critical role in supporting adaptive behavior; however, whether it can be causally regulated in humans via deep subcortical stimulation remains unclear. In the present study, we investigated the differential effects of substantia nigra (SN) and subthalamic nucleus (STN) stimulation on episodic memory, as well as the underlying mechanisms of its associated brain networks, using a recognition memory task combined with concurrent functional magnetic resonance imaging in patients with Parkinson’s disease. SN-DBS increased recognition sensitivity and reduced false alarms at both frequencies, whereas 10 Hz STN-DBS reduced sensitivity and increased false alarms. Functional connectivity analyses in the absence of DBS stimulation identified a false recognition-related network linking nigral, pallidal, subthalamic, medial temporal, frontal, and occipital regions. SN-DBS-related false alarm reduction tracked modulation of this circuit and was marked by its baseline vulnerability state. These behavioral effects mapped onto target-dependent parieto-occipital and SN–visual retrieval pathways, supporting a model in which DBS bidirectionally regulates recognition memory through target- and frequency-dependent subcortical–cortical circuits.

## Introduction

One of the brain’s most adaptive capacities is to recognize when present experience echoes the past. A familiar scene can guide behavior, preserve continuity of the self and allow previous experience to shape future choice; yet the same capacity can also keep fear, stress or grief accessible long after they cease to be useful^1^. Episodic memory is therefore not a passive archive but a dynamic trace that binds items to their spatial, temporal and internal context and reconstructs those events when they are needed^2, 3^. Effective control of episodic memory must consequently be bidirectional: it may require strengthening weak but useful memories, while also suppressing, attenuating or remodeling intrusive memories whose emotional, fearful or stress-related content has become maladaptive^4–7^.

In model organisms, the mechanisms supporting episodic-like memory have become increasingly tractable. Hippocampal–prefrontal interactions contribute to the integration of what happened with where and when it occurred, and direct manipulation of engram-bearing ensembles has established that specific memory traces can be reactivated and reassigned at the circuit level^2,7–9^. At the same time, memory-relevant processing is not confined to the hippocampal formation. Recent work has shown that cortico–basal ganglia–thalamic loops are organized as structured, recurrent subnetworks, and that the thalamus actively coordinates functional interactions within and across cortical territories^10, 11^. Importantly, deep dopaminergic signals arising from the ventral tegmental area and substantia nigra can directly facilitate associative encoding in entorhinal cortex, demonstrating in mechanistically explicit terms that interventions at subcortical nuclei can influence cortical memory machinery^12^. A specific role in mesolimbic-hippocampal interplay has been proposed to determine the encoding of novel stimuli into episodic memory^13^. These findings raise a central translational possibility: deep brain neuromodulation may regulate memory not only by acting locally in subcortex, but by reshaping distributed cortical networks that support remembering.

Yet these advances also expose a translational gap. Animal experiments offer causal precision, but they necessarily rely on simplified, experimenter-defined behaviors and cannot fully capture the autobiographical richness and social embedding of human episodic memory^1–3^. Whether deep subcortical circuits can be used to causally modulate human episodic memory therefore remains unresolved. Existing human evidence is informative but sparse: reactivated episodic memories can be disrupted under specific conditions, and direct stimulation of medial temporal or cortical memory circuits can either improve or impair performance depending on timing, target and stimulation strategy^14–17^. Thus, human literature supports the principle that memory is modifiable, but it not yet established how targeting deep nuclei might be used to regulate episodic memory in a circuit-specific manner.

Parkinson’s disease (PD) offers a rare opportunity to address this problem in an ethical and mechanistically informative human model. Deep brain stimulation (DBS) is an established therapy for medication-refractory PD and provides controlled access to deep basal ganglia/diencephalon and mesolimbic nuclei in awake, behaving patients^18–20^. This approach to studying the functional role of these deep structures in memory is particularly relevant because cognitive impairment is common in PD, memory is frequently affected, and current DBS practice is optimized primarily for motor rather than cognitive goals^18, 19^. In this setting, DBS is not merely a therapeutic tool; it becomes a means of causally testing how subcortical nodes participate in human memory networks.

Within this framework, we focused on two neighboring but circuit-distinct targets: the substantia nigra (SN) and the subthalamic nucleus (STN). The SN was selected as a candidate pro-mnemonic target because it sits at the interface of salience, dopamine and memory. In animal work, dopamine signals from the ventral tegmental area/SN facilitate associative encoding in entorhinal cortex and promote incorporation of new associations into an existing cognitive map^12^. In humans, intraoperative single-neuron recordings have identified putative dopaminergic SN neurons that discriminate novelty from familiar stimuli, update rapidly after minimal exposure and synchronize with frontal theta activity in proportion to later declarative memory success^21^. More recent single-neuron evidence further suggests that human SN neurons are modulated by cognitive boundaries during ongoing experience, and that a subset of boundary-responsive neurons differentiates novel from familiar images during recognition and predicts memory success. Together, these findings make the SN a mechanistically grounded target for testing whether stimulation can enhance recognition memory and recruit distributed cortical systems during retrieval.

The STN was selected for a complementary reason. It was not chosen as a priori memory-enhancing target, but as a clinically essential and mechanistically informative comparative target. The STN is the canonical DBS target in PD, yet it is not simply a motor relay: it receives direct frontal input, participates in hyperdirect circuits for action selection and cognitive control, and is positioned to reshape large-scale frontal and thalamo-cortical interactions when stimulated^19–24^. Because episodic retrieval depends not only on stored representations but also on control processes that select, evaluate and reject competing traces, STN stimulation could plausibly influence recognition performance either by supporting cognitive control-related computations or by perturbing them. Comparing SN and STN stimulation therefore allows a direct test of whether target identity within closely related cortico-subcortical loops determines the direction of memory modulation.

Frequency was treated as a second critical axis of causality. We compared 10 Hz and 130 Hz stimulation because these frequencies are expected to engage distinct physiological regimes. High-frequency 130 Hz stimulation represents the conventional therapeutic frequency for PD and is closely linked to suppression of pathological beta synchrony and motor improvement18, 23. By contrast, 10 Hz was chosen as a mechanistically informed low-frequency condition because episodic memory and cognitive control are strongly coupled to theta-range dynamics, including low-frequency hippocampal–cortical and fronto-subthalamic interactions, and because prior STN studies indicated that stimulation near this range can modulate episodic-category fluency and working memory^24, 26–28^. Rather than assuming that one frequency would be uniformly beneficial across targets, we reasoned that low- and high-frequency DBS might differentially bias memory-relevant networks.

Here, in a single-blind, randomized, prospective study of patients with PD, we compared SN-DBS and STN-DBS during a recognition-memory paradigm while combining behavioral testing with concurrent resting-state and event-related fMRI across OFF, 10 Hz and 130 Hz stimulation conditions. This design allowed us to ask whether deep target and stimulation frequency jointly determine memory outcome, and whether any behavioral dissociation can be explained by reconfiguration of cortico-subcortical network interactions. By doing so, we sought to move the study of episodic memory regulation from descriptive association toward target-specific causal circuit modulation in humans.

## Results

### Cohort characteristics and target localization

In this study, a total of 33 patients with PD scheduled for bilateral STN-DBS implantation were enrolled (mean age: 60.79 ± 7.17 years; 23 males; mean PD duration: 9.24 ± 2.96 years). Within an ethically approved prospective experimental framework, participants were randomly allocated in equal numbers to one of two predefined implantation strategies: an SN-DBS group or an STN-DBS group. In the SN-DBS group, the DBS lead trajectory was designed to span the STN and extend into the SN, typically leaving two to three contacts within the STN for standard clinical programming and the most ventral contact within the SN for experimental stimulation. In the SN-DBS group (n = 17), electrode implantation was performed such that the deepest DBS contact was positioned within substantia nigra pars compacta/reticulata. Given the spatial resolution of BOLD fMRI, we refer to the stimulated region encompassing both substantia nigra pars compacta and pars reticulata as the SN and interpret the findings at the network level. In the STN-DBS group (n = 16), the deepest DBS contact was positioned in the ventral subregion of the STN. (***Supplementary Material Figure 1***). Approximately one month after DBS implantation, patients underwent postoperative computed tomography (CT) to verify lead localization and then completed three separate MRI sessions in the clinically defined medication-off, each acquired under one stimulation condition (OFF, 10 Hz, or 130 Hz; stimulation order was randomized across participants). Hereafter, OFF refers specifically to the DBS stimulation that was turned off. Before each MRI acquisition, any change in stimulation setting was followed by a washout/stabilization interval of at least 30 min to minimize carry-over effects between conditions. Each session included a T1-weighted structural scan, resting-state fMRI and event-related fMRI (**Fig.1 a**). For participants in the SN-DBS group, the volume of tissue activated by DBS was constrained to remain predominantly within the SN. This was ensured by performing individualized electrode reconstruction and the volume of tissue activated modeling in Lead-DBS v3.2^29^ (https://www.lead-dbs.org) before experimental testing. For both groups, candidate stimulation settings were simulated in advance, and only parameters producing a model of the volume of tissue activated confined to the intended target region were used during the experiment. (**Fig.1 b, c**). An additional 16 age-matched healthy control (HC) participants were also enrolled and completed a single MRI session for cross-sectional comparison.

**Fig. 1.**
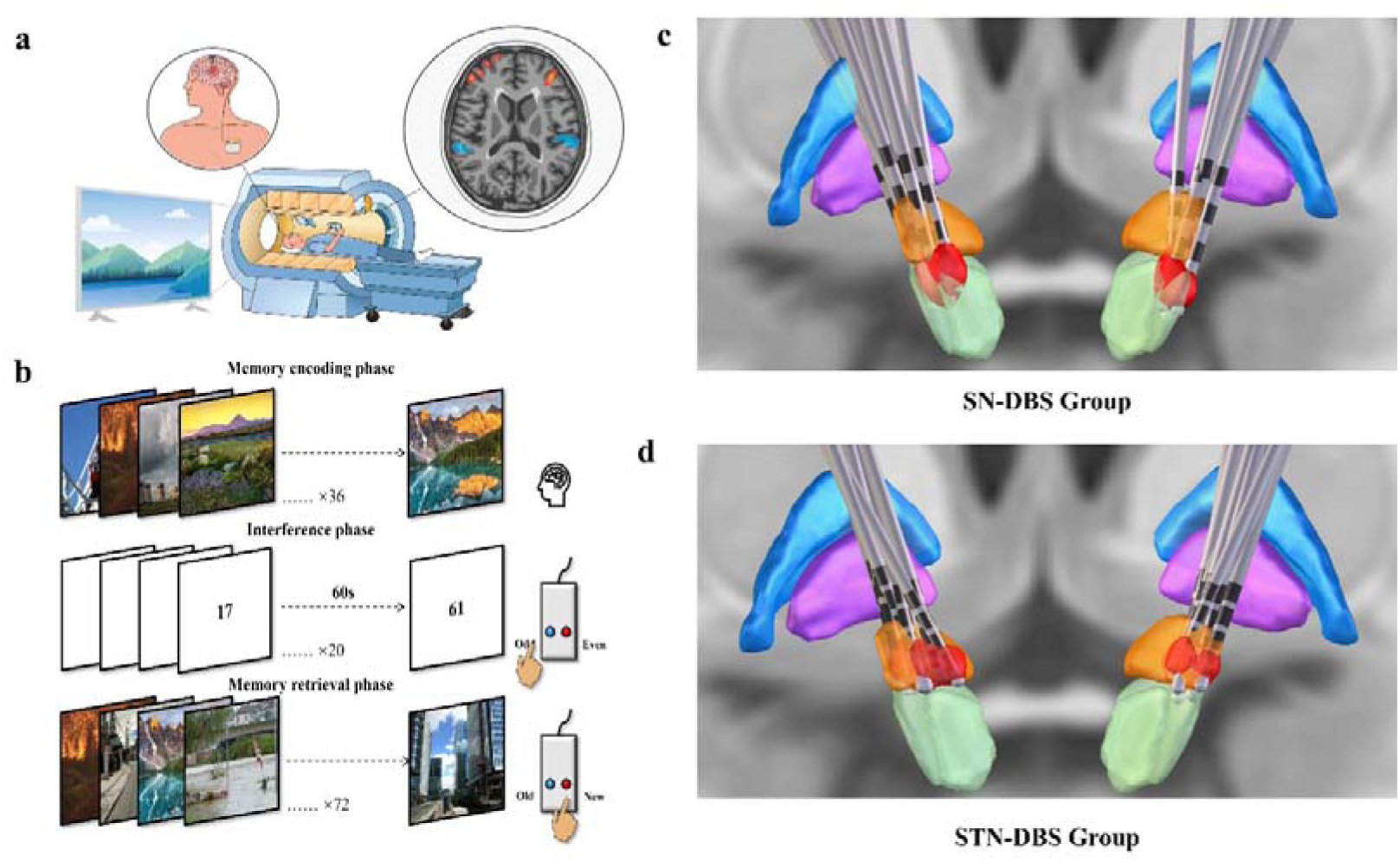
Experimental procedure and participant grouping. **(a)** Schematic illustration of the event-related fMRI experimental setup; **(b)** Schematic illustration of the episodic memory task. The paradigm consisted of three phases: during the encoding phase, participants were instructed to memorize 36 scene-related images; during the interference phase, participants performed a digit parity judgment task and responded via button press; during the retrieval phase, participants were asked to determine whether each of 72 images had been previously presented during the encoding phase and to respond as quickly and accurately as possible. The images used in the task were visually similar but not identical, requiring fine-grained memory recognition discrimination; **(c)** DBS lead implantation sites and the volume of tissue activated distribution across all participants in the SN-DBS group. The globus pallidus externus (GPe) is shown in blue, the globus pallidus internus (GPi) in purple, the STN in orange, the SN (including pars compacta and pars reticulata) in green; the volume of tissue activated is shown in red. **(d)** DBS lead implantation sites and the volume of tissue activated distribution across all participants in the STN-DBS group.

Participants completed an event-related old/new recognition-memory task during fMRI, requiring discrimination of studied images from perceptually similar foils (**Fig.1 d**; see Methods). Behavioral performance was summarized using recognition accuracy, signal-detection measures and response times.

### Motor improvement was restricted to 130 Hz STN-DBS

UPDRS-III scores were obtained under each stimulation condition after parameter adjustment and a stabilization period before MRI acquisition. In the STN-DBS group, UPDRS-III scores differed significantly across stimulation conditions (p < 0.001). Post hoc paired comparisons showed that 130 Hz STN-DBS reduced UPDRS-III scores relative to both OFF stimulation (28.25 ± 17.07 vs. 38.25 ± 18.17; q = 0.011) and 10 Hz stimulation (36.63 ± 20.80; q = 0.004), whereas 10 Hz stimulation did not differ from OFF. In contrast, UPDRS-III scores did not differ significantly across stimulation conditions in the SN-DBS group (p = 0.078), and no FDR-corrected pairwise comparison was significant. Thus, motor improvement was restricted to 130 Hz STN-DBS. (***Supplementary Materials Table 1***).

### SN-DBS and STN-DBS exerted opposing effects on recognition sensitivity

Participants completed an old/new recognition-memory task in which previously encoded images and visually similar novel foils were judged as old or new^30^ (**Fig.2 a**). Retrieval responses were classified as hits, misses, false alarms or correct rejections, and signal-detection measures were calculated after excluding trials with response times <200 ms or no response. Before examining DBS-induced changes, we compared baseline recognition performance between HCs and PD patients under OFF stimulation using Welch’s tests with FDR correction. Relative to HCs, PD patients under OFF stimulation showed poorer recognition-memory performance, reflected by lower d′ and Pr, higher false alarm rate, lower hit rate, slower correct-rejection responses and a higher no-response rate, whereas response bias c and hit reaction time did not survive correction. Baseline performance did not differ reliably between the SN-OFF and STN-OFF subgroups, supporting the subsequent within-target DBS comparisons (***Supplementary Materials Table 4***).

**Fig. 2.**
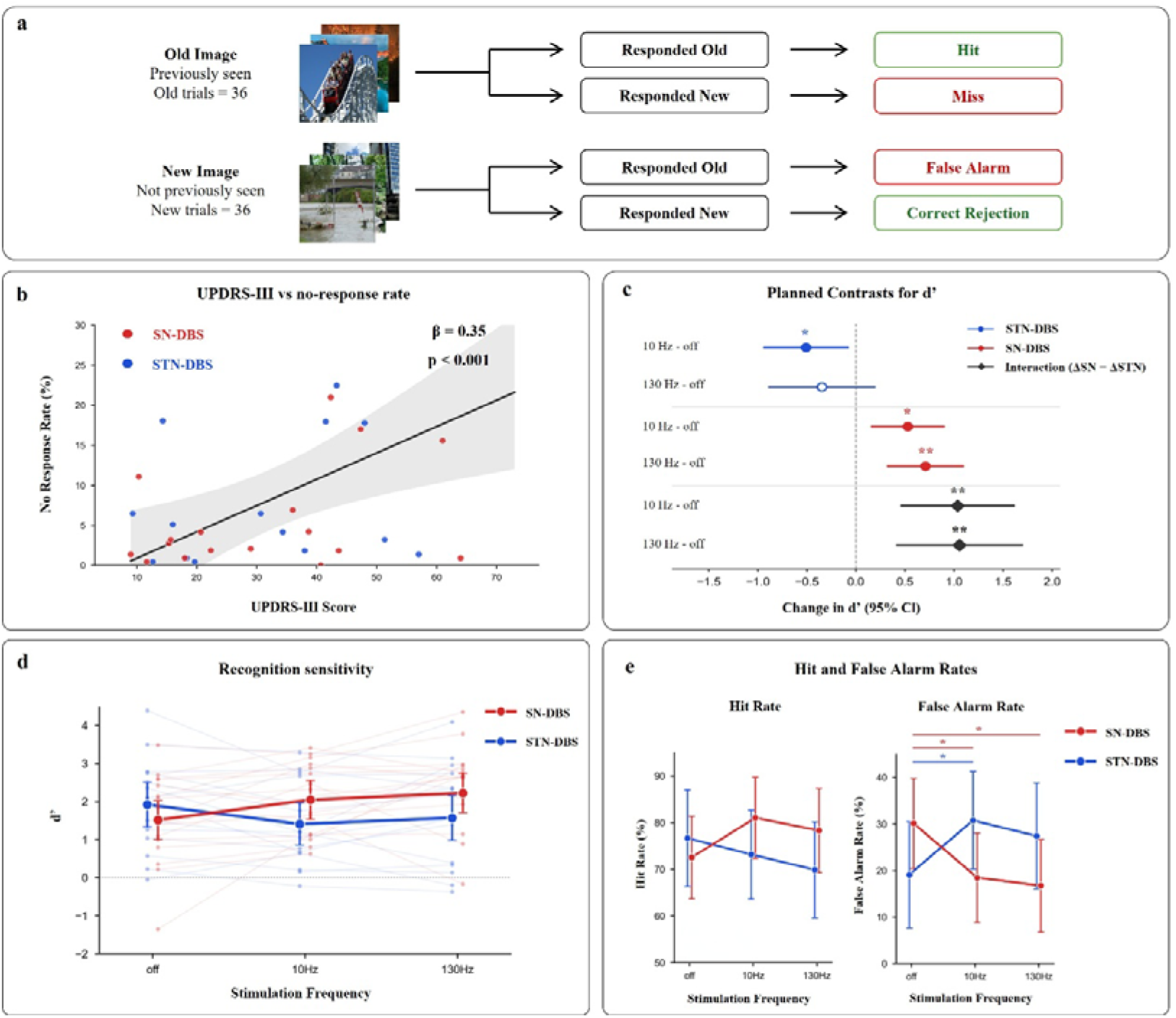
Target-dependent DBS effects on recognition memory after accounting for motor status. **(a)** Schematic classification of recognition-memory retrieval outcomes. Previously encoded images judged as old were classified as hits, whereas previously encoded images judged as new were classified as misses. Novel images judged as old were classified as false alarms, whereas novel images judged as new were classified as correct rejections. Trials with response times < 200 ms or no response were excluded from analyses. **(b)** Relationship between motor severity and no-response rate. Each point represents one participant averaged across retained stimulation sessions. The regression line and shaded band are shown for visualization. Mean UPDRS-III score was positively associated with no-response rate, indicating that motor status affected task omissions and motivating UPDRS-III adjustment in the behavioral models. **(c)** Contrasts for recognition sensitivity, indexed by d′, from the UPDRS-III-adjusted mixed-effects model. The upper four rows show within-target stimulation effects relative to OFF, defined as DBS-active − OFF, separately for STN-DBS and SN-DBS. The lower two rows show target-interaction contrasts, defined as ΔSN − ΔSTN, where Δ denotes DBS-active − OFF. Positive interaction estimates therefore indicate a larger stimulation-related increase in d′ under SN-DBS than under STN-DBS. Points indicate model-estimated contrasts and horizontal bars indicate 95% confidence intervals. Asterisks denote FDR-corrected contrasts. **(d)** Recognition sensitivity across stimulation conditions. Pale lines and small points indicate raw d′ trajectories. Large colored points and error bars indicate UPDRS-III-adjusted model-estimated marginal means and 95% confidence intervals. **(e)** Hit rate and false alarm rate from the UPDRS-III-adjusted mixed-effects models. Error bars indicate 95% confidence intervals. Hit rate did not show FDR-corrected stimulation effects, whereas false alarm rate showed target-dependent modulation: SN-DBS reduced false alarms at both 10 Hz and 130 Hz, whereas 10 Hz STN-DBS increased false alarms. Red denotes SN-DBS and blue denotes STN-DBS. Behavioral mixed-effects models included stimulation target, stimulation frequency, their interaction, between-subject UPDRS-III, within-subject UPDRS-III and a subject-level random intercept. Contrasts were corrected using the Benjamini–Hochberg false discovery rate procedure.

Because DBS manipulations can alter motor state, we first examined whether motor severity affected task engagement. Mean UPDRS-III score was positively associated with no-response rate (β = 0.35, 95% CI [0.15, 0.54], p < 0.001; **Fig.2 b**). This indicated that motor status influenced task omissions. Therefore, all primary behavioral models included UPDRS-III score as a covariate, decomposed into between-subject and within subject components, together with stimulation target, stimulation frequency, their interaction and a subject level random intercept.

Recognition sensitivity, indexed by d′, showed target-dependent stimulation modulation after adjustment for UPDRS-III score. Raw within-subject trajectories and UPDRS-III adjusted model estimates showed that d′ increased under SN-DBS but decreased or failed to improve under STN-DBS, particularly at 10 Hz (**Fig.2 c, d**). Contrasts from the adjusted mixed effects model confirmed this pattern. In the STN-DBS group, 10 Hz stimulation significantly reduced d′ relative to OFF (β = −0.51, 95% CI [−0.94, −0.08], q = 0.036), whereas 130 Hz stimulation did not reach significance (β = −0.35, 95% CI [−0.89, 0.20], q = 0.236). By contrast, in the SN-DBS group, d′ increased significantly at both 10 Hz (β = 0.53, 95% CI [0.16, 0.90], q = 0.011) and 130 Hz (β = 0.71, 95% CI [0.33, 1.10], q = 0.002) relative to OFF.

The target-dependent interaction was significant at both stimulation frequencies. At 10 Hz, the difference in stimulation induced change between targets, defined as ΔSN − ΔSTN, was β = 1.04 (95% CI [0.46, 1.61], q = 0.002). At 130 Hz, the corresponding interaction was β = 1.06 (95% CI [0.42, 1.69], q = 0.003). Thus, SN-DBS and STN-DBS produced bidirectional effects on recognition sensitivity even after accounting for motor-state-related task omissions.

We next examined whether the d′ effect was driven by changes in hit rate or false alarm rate. Hit rate (number of correct hits divided by the number of valid old image trials) showed the expected directional pattern, with numerical increases under SN-DBS and decreases under STN-DBS, but no planned hit-rate contrast survived FDR correction (Fig.2 e). In contrast, false alarm rate (number of false alarms divided by the number of valid new image trials) showed robust target-dependent modulation. In the STN-DBS group, 10 Hz stimulation significantly increased false alarm rate relative to OFF (Δ = +11.7 percentage points, 95% CI [+1.6, +21.8], q = 0.042). Conversely, in the SN-DBS group, false alarm rate was significantly reduced at both 10 Hz (Δ = −11.7 percentage points, 95% CI [−20.3, −3.0], q = 0.018) and 130 Hz (Δ = −13.4 percentage points, 95% CI [−22.4, −4.4], q = 0.014).

The target-dependent false-alarm interaction was significant at both frequencies. At 10 Hz, the interaction was −23.4 percentage points (95% CI [−36.9, −9.8], q = 0.006). At 130 Hz, the interaction was −21.7 percentage points (95% CI [−36.8, −6.7], q = 0.014). These findings indicate that the memory benefit of SN-DBS was primarily expressed as reduced false recognition of novel but visually similar images, whereas 10 Hz STN-DBS increased false recognition. All q values reported in this section represent Benjamini–Hochberg FDR-adjusted p values. Note that correct rejection rate of new stimuli during recognition is (1 – false alarm rate); we focus on the false alarm rate in what follows because it is used in the calculation of d’.

Secondary behavioral analyses are provided in the Supplementary Information. Pr closely paralleled the d′ findings, whereas response bias c, hit reaction time and correct-rejection reaction time did not show corresponding stimulation-related effects. Subject-level change-score heatmaps and individual Δd′ trajectories further illustrated interindividual variability and confirmed that the target-dependent behavioral pattern was not driven by a single outlier (***Supplementary Fig. 5***).

Having established that SN-DBS and STN-DBS produced opposing effects on recognition memory, with false alarm rate providing the clearest behavioral signature of target-dependent modulation, we next provide a mechanistic basis for this observation, drawing on resting-state fMRI followed by task-evoked fMRI responses during recognition testing.

### Baseline functional connectivity of deep-brain structures relates to recognition performance

We analysed data from each patient’s resting-state fMRI session to determine whether individual differences in recognition performance were related in intrinsic functional connectivity of disease- and memory-relevant deep-brain circuits under OFF. We selected bilateral seed regions a priori from subcortical and medial temporal masks. The core PD/DBS seed set included SN, STN, GPi, GPe, putamen and thalamic/VIM-related regions, motivated by recent evidence that the SN and canonical PD DBS targets are selectively connected with the somato-cognitive action network (SCAN)^31^. We further included hippocampus and parahippocampal regions as medial temporal structures implicated in novelty, familiarity, mnemonic evidence and recognition-memory decisions. Deep nuclei were defined primarily using HybraPD^32^ and DISTAL atlas masks^33^, supplemented with AAL-90^34^ anatomical labels for medial temporal regions.

We first performed OFF seed-to-voxel functional connectivity analyses during OFF and related baseline connectivity to recognition sensitivity, indexed by d′. Baseline d′ was associated with three circumscribed connectivity patterns (**Fig.3 a**). Lower d′ was associated with stronger right GPi connectivity to a right middle/inferior frontal cluster and stronger right nucleus accumbens connectivity to a right middle/superior frontal cluster. Conversely, higher d′ was associated with stronger right GPi connectivity to a precuneus cluster. Thus, baseline recognition sensitivity in PD patients was related to pallidal and accumbens coupling with frontal and posterior cortical regions.

**Fig. 3.**
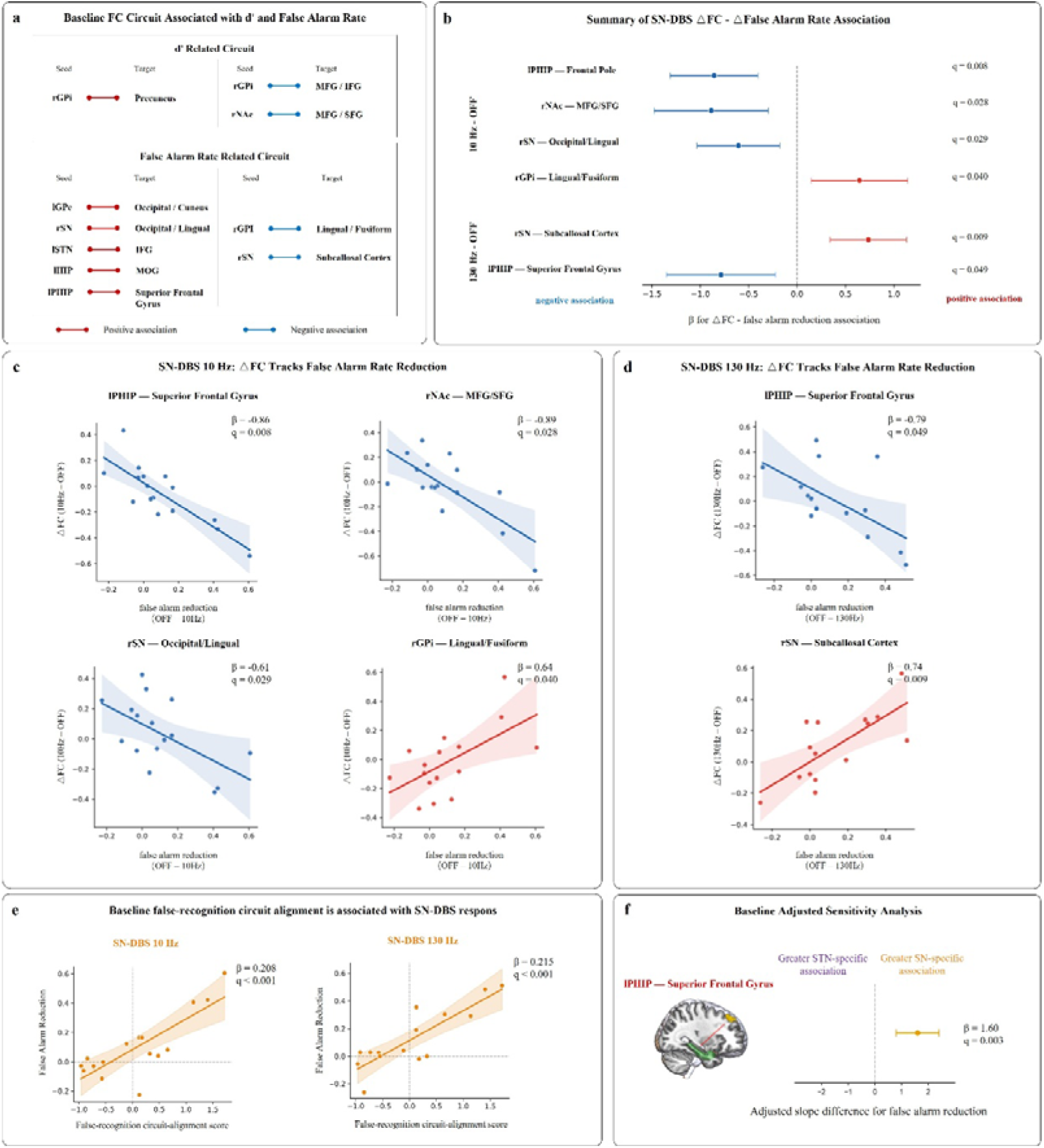
OFF false-recognition circuit and the reverse modulation by SN-DBS. **(a)** Baseline FC circuits associated with d′ and false alarm rate under OFF. For d′, higher recognition sensitivity was associated with stronger right GPi–precuneus connectivity, whereas lower d′ was associated with stronger right GPi–middle/inferior frontal and right nucleus accumbens–middle/superior frontal connectivity. For false alarm rate, higher false alarm rate was associated with stronger left GPe–occipital/cuneus, right SN–occipital/lingual, left STN–inferior frontal gyrus (IFG), left hippocampus–middle occipital gyrus (MOG), and left parahippocampal gyrus (PHG)–superior frontal gyrus connectivity, whereas lower false alarm rate was associated with stronger right GPi–lingual/fusiform and right SN–subcallosal cortex connectivity. Red lines indicate positive associations and blue lines indicate negative associations. **(b)** Summary of significant associations between SN-DBS-induced connectivity change and false alarm reduction. At 10 Hz SN-DBS, greater false alarm reduction was associated with decreased left PHG–superior frontal gyrus connectivity, decreased right nucleus accumbens–middle/superior frontal connectivity, decreased right SN–occipital/lingual connectivity, and increased right GPi–lingual/fusiform connectivity. At 130 Hz SN-DBS, greater false alarm reduction was associated with decreased left PHG–superior frontal gyrus connectivity and increased right SN–subcallosal connectivity. The direction of these effects was reversed to the OFF false alarm map: connections positively associated with false alarm under OFF tended to decrease, whereas connections negatively associated with false alarm under OFF tended to increase. **(c)** At 10 Hz SN-DBS, FC changes tracked false alarm reduction. **(d)** At 130 Hz SN-DBS, FC changes also tracked false alarm reduction. **(e)** Baseline false-recognition circuit-alignment score was associated with false alarm reduction during SN-DBS at both 10 Hz and 130 Hz. Higher scores indicate stronger alignment with the OFF connectivity pattern associated with higher false alarm rate. Each point represents one participant. **(f)** Baseline-adjusted sensitivity analysis identified L-PHG–superior frontal gyrus connectivity as a target-dependent marker at 10 Hz. The plotted estimate represents the adjusted SN − STN slope difference; positive values indicate a stronger association under SN-DBS than STN-DBS. q values indicate FDR-adjusted p values.

Because the behavioral benefit of SN-DBS was primarily driven by reduced false recognition of novel images, we next focused on OFF connectivity associated with false alarm rate. This analysis identified a broader false-recognition circuit (**Fig.3 a**). False alarm rate was positively associated with left GPe–occipital/cuneal connectivity, right SN–occipital/lingual connectivity, left STN–inferior frontal connectivity, left hippocampal–middle occipital connectivity and left parahippocampal–superior frontal gyrus connectivity. False alarm rate was negatively associated with right GPi–lingual/fusiform connectivity and right SN–subcallosal connectivity. These findings indicate that false recognition under OFF was not linked to a single region, but to intrinsic coupling among basal ganglia and nigral seeds, medial temporal structures, frontal cortex and posterior visual areas.

### SN-DBS-related false alarm reduction tracks modulation of the OFF false-recognition circuit

We next asked whether behavioral improvement was accompanied by modulation of the same circuit identified under OFF. For each OFF false alarm-related seed–cluster pair, we extracted Fisher-z connectivity under OFF, 10 Hz and 130 Hz. Stimulation-induced connectivity change was computed as ΔFC = FC (DBS-active) – FC (OFF). Behavioral improvement was indexed by false alarm reduction, defined as false alarm rate (OFF) − false alarm rate (DBS-active), such that positive values indicate fewer false alarms during active stimulation. A summary of the significant SN-DBS-related ΔFC–behavior associations is shown in **Fig.3 b**. At 10 Hz SN-DBS, greater false alarm reduction was associated with decreased left PHG–superior frontal gyrus connectivity (β = −0.86, q = 0.008), decreased right nucleus accumbens–middle/superior frontal connectivity (β = −0.89, q = 0.028), decreased right SN–occipital/lingual connectivity (β = −0.61, q = 0.029), and increased right GPi–lingual/fusiform connectivity (β = 0.64, q = 0.040; **Fig.3 c**). At 130 Hz SN-DBS, greater false alarm reduction was associated with decreased left PHG–superior frontal gyrus connectivity (β = −0.79, q = 0.049) and increased right SN–subcallosal connectivity (β = 0.74, q = 0.009; **Fig.3 d**).

The directions of these effects were systematically opposite to the OFF false alarm map. Connections positively associated with false alarm rate under OFF tended to decrease in participants showing greater false alarm reduction during SN-DBS, whereas connections negatively associated with false alarm rate under OFF tended to increase. Thus, SN-DBS did not produce a uniform increase or decrease in connectivity. Rather, behavioral benefit tracked a reverse modulation of the pre-existing OFF false-recognition circuit. No corresponding STN-DBS ΔFC–false alarm reduction association survived correction. Notably, left PHG–superior frontal gyrus connectivity was implicated at both stimulation frequencies, suggesting that medial temporal–frontal coupling may represent a shared circuit component of SN-DBS-related reduction in false-recognition.

### Baseline false-recognition circuit alignment was associated with SN-DBS response

We then asked whether the baseline configuration of the OFF false-recognition circuit was associated with the magnitude of subsequent SN-DBS benefit. To quantify each participant’s baseline circuit state, we computed a sign-oriented false-recognition circuit-alignment score from the OFF false alarm-related connections. Connections positively associated with false alarm rate under OFF were entered with a positive sign, whereas connections negatively associated with false alarm rate under OFF were sign-reversed. The oriented values were z-scored and averaged across connections, such that higher scores indicated stronger alignment with the OFF connectivity pattern associated with higher false recognition.

Higher baseline false-recognition circuit-alignment scores were associated with greater false alarm reduction during SN-DBS at both 10 Hz (β = 0.208, q < 0.001) and 130 Hz (β = 0.215, q < 0.001; **Fig. 3e**). Direct slope comparisons showed that this alignment–response relationship was stronger for SN-DBS than for STN-DBS at both frequencies, supporting target specificity. These findings indicate that participants whose baseline connectivity more closely resembled the false-recognition-related OFF circuit tended to show larger SN-DBS- related behavioral benefit.

However, because false alarm reduction includes baseline false alarm rate, this analysis was interpreted as a baseline circuit-marker analysis rather than an independent prediction model. We therefore performed baseline-adjusted sensitivity analyses. Most composite alignment effects were attenuated after adjustment for baseline false alarm rate, indicating that the composite score primarily indexed baseline false-recognition liability. However, left parahippocampal–superior frontal gyrus connectivity retained a target-dependent effect after adjustment at 10 Hz (adjusted SN − STN slope difference: β = 1.60, 95% CI [0.80, 2.40], q = 0.003; **Fig. 3f**). This result identifies the parahippocampal–superior frontal gyrus connection as the strongest adjusted baseline marker distinguishing SN-DBS and STN-DBS effects on false alarm reduction.

### ALFF changes reveal target-dependent parieto-occipital modulation of false recognition

The preceding analyses showed that SN-DBS-related false alarm reduction was associated with modulation of an OFF false-recognition connectivity circuit. We next asked whether the same behavioral effect was accompanied by changes in spontaneous local activity. We therefore performed whole-brain analyses of amplitude of low-frequency fluctuation (ALFF), focusing on stimulation-induced ALFF changes and their relationship with false alarm reduction. ΔALFF was defined as ALFF (DBS-active) – ALFF (OFF), and false alarm reduction was defined as false alarm rate (OFF) − false alarm rate (DBS-active), such that positive false alarm reduction indicates reduced false recognition during active DBS. The primary ALFF model tested group-specific behavioral slopes of ΔALFF against false alarm reduction, together with the SN-DBS versus STN-DBS slope difference.

At 10 Hz, SN-DBS and STN-DBS showed opposite ALFF–behavior relationships. In the SN-DBS group, greater false alarm reduction was associated with increased ΔALFF in dorsal parieto-occipital and somatosensory regions, including left superior parietal/lateral occipital cortex, bilateral postcentral/anterior supramarginal regions and left lateral occipital/precuneus cortex (**Fig.4 a**). ROI-level extraction from these clusters confirmed positive SN-DBS slopes, including the representative left superior parietal/lateral occipital cluster shown in **Fig.4 b** (β = 0.38, q < 0.001). Thus, in participants receiving SN-DBS, greater reduction of false recognition at 10 Hz was accompanied by increased low-frequency amplitude in a dorsal parieto-occipital network.

**Fig. 4.**
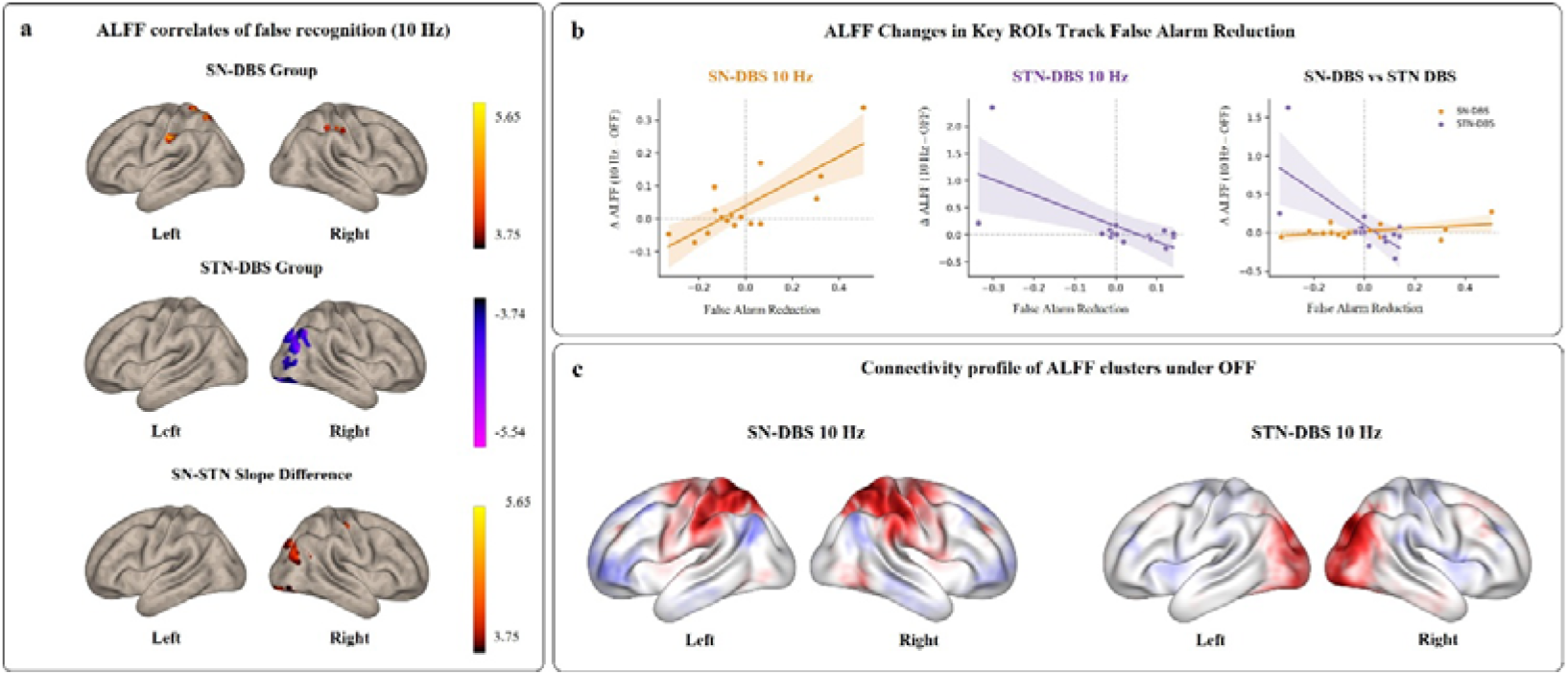
Target-dependent ALFF correlates of false alarm reduction at 10 Hz. **(a)** Whole-brain ALFF analyses showing regions in which stimulation-induced ALFF changes were associated with false alarm reduction at 10 Hz. ΔALFF was defined as ALFF (10 Hz) – ALFF (OFF). False alarm reduction was defined as false alarm reduction = false alarm rate (OFF) − false alarm rate (10 Hz). In the SN-DBS group, greater false alarm reduction was associated with increased ΔALFF in dorsal parieto-occipital and postcentral regions. In the STN-DBS group, greater false alarm reduction was associated with decreased ΔALFF in right occipital visual regions. Direct SN-DBS versus STN-DBS slope comparisons identified target-dependent ALFF–false alarm reduction effects in right occipital and precentral/postcentral regions. All imaging results were thresholded at p < 0.001 at the voxel level, with cluster-level FWE correction at p < 0.05. **(b)** ROI-level effect-size visualization from representative ALFF clusters. Left, SN-DBS at 10 Hz showed a positive relationship between false alarm reduction and ΔALFF in a left superior parietal/lateral occipital cluster. Middle, STN-DBS at 10 Hz showed a negative relationship between false alarm reduction and ΔALFF in a right superior lateral occipital/occipital pole cluster. Right, representative target-difference cluster showing opposite SN-DBS and STN-DBS slopes. ROI extraction was used for visualization and effect-size estimation. **(c)** Average OFF connectivity profiles of representative ALFF clusters. The SN-DBS-related ALFF cluster was embedded in a bilateral dorsal parieto-occipital/sensorimotor connectivity pattern, whereas the STN-DBS-related visual cluster showed posterior visual connectivity with additional sensorimotor involvement. These maps contextualize the ALFF clusters within broader sensorimotor–visual networks. q values are FDR-corrected within the model × frequency × outcome × test family.

In contrast, the STN-DBS group showed the opposite pattern. Greater false alarm reduction was associated with decreased ΔALFF in right visual regions, including superior lateral occipital cortex/occipital pole, inferior lateral occipital cortex/occipital fusiform cortex/occipital pole and inferior/superior lateral occipital cortex (**Fig.4 a**). ROI extraction from the representative right superior lateral occipital/occipital pole cluster showed a significant negative STN-DBS slope (β = −2.85, q < 0.001; **Fig.4 b**). This indicates that 10 Hz STN-DBS participants with less false alarm reduction, or greater false recognition, tended to show stronger ALFF increases in right occipital visual cortex.

Direct comparison of SN-DBS and STN-DBS slopes confirmed target-dependent ALFF–behavior coupling at 10 Hz. Significant slope differences were observed in right lateral occipital/occipital pole, right occipital fusiform/inferior lateral occipital regions and right precentral/postcentral cortex (**Fig.4 a**). In the representative right lateral occipital/occipital pole cluster, the slope difference was driven by a negative STN-DBS relationship and a weaker positive SN-DBS tendency (β = 2.39, q < 0.001; **Fig.4 b**). These results indicate that the local ALFF correlates of false recognition were target dependent: SN-DBS recruited dorsal parieto-occipital and postcentral regions in the direction of behavioral improvement, whereas STN-DBS showed an opposing relationship in right occipital visual cortex.

No comparable whole-brain ALFF–false alarm rate association was detected at 130 Hz. Thus, although SN-DBS reduced false alarm rate at both 10 Hz and 130 Hz behaviorally, the ALFF signature of false alarm reduction was most apparent at 10 Hz. Together, these findings provide convergent regional evidence that target-dependent modulation of false recognition is accompanied by changes in local low-frequency amplitude within parieto-occipital, visual and sensorimotor systems.

### Target-dependent SN–visual coupling during memory retrieval

We next asked whether the DBS target-dependent behavioral effects were accompanied by task-evoked changes in functional coupling during memory retrieval. We constructed event-related GLMs using the onset time series of old and new images during the retrieval stage and then performed generalized psychophysiological interaction (gPPI) analyses. Retrieval-stage quality check was applied before gPPI modelling, and participants or sessions with excessive missing responses or insufficient valid retrieval events were excluded from this analysis.

The gPPI analysis used priori ROI templates spanning deep-brain, medial temporal and cortical systems, as defined in the Methods. The subcortical and medial temporal seed set included GPe, putamen, GPi, STN, nucleus accumbens, SN, hippocampus, parahippocampal cortex and amygdala. Cortical ROIs were grouped from AAL-90 anatomical labels into functional systems, including DLPFC, inferior frontal, orbitofrontal, medial frontal, motor/sensorimotor, insula, cingulate, parietal, precuneus, visual occipital, fusiform and temporal systems. In the main event-related fMRI analysis, we focused on two gPPI measures: new-image coupling, which indexes connectivity during presentation of novel retrieval items, and old minus new (old – new) coupling, which indexes differential connectivity during old versus new image processing (**Fig.5 a**).

**Fig. 5.**
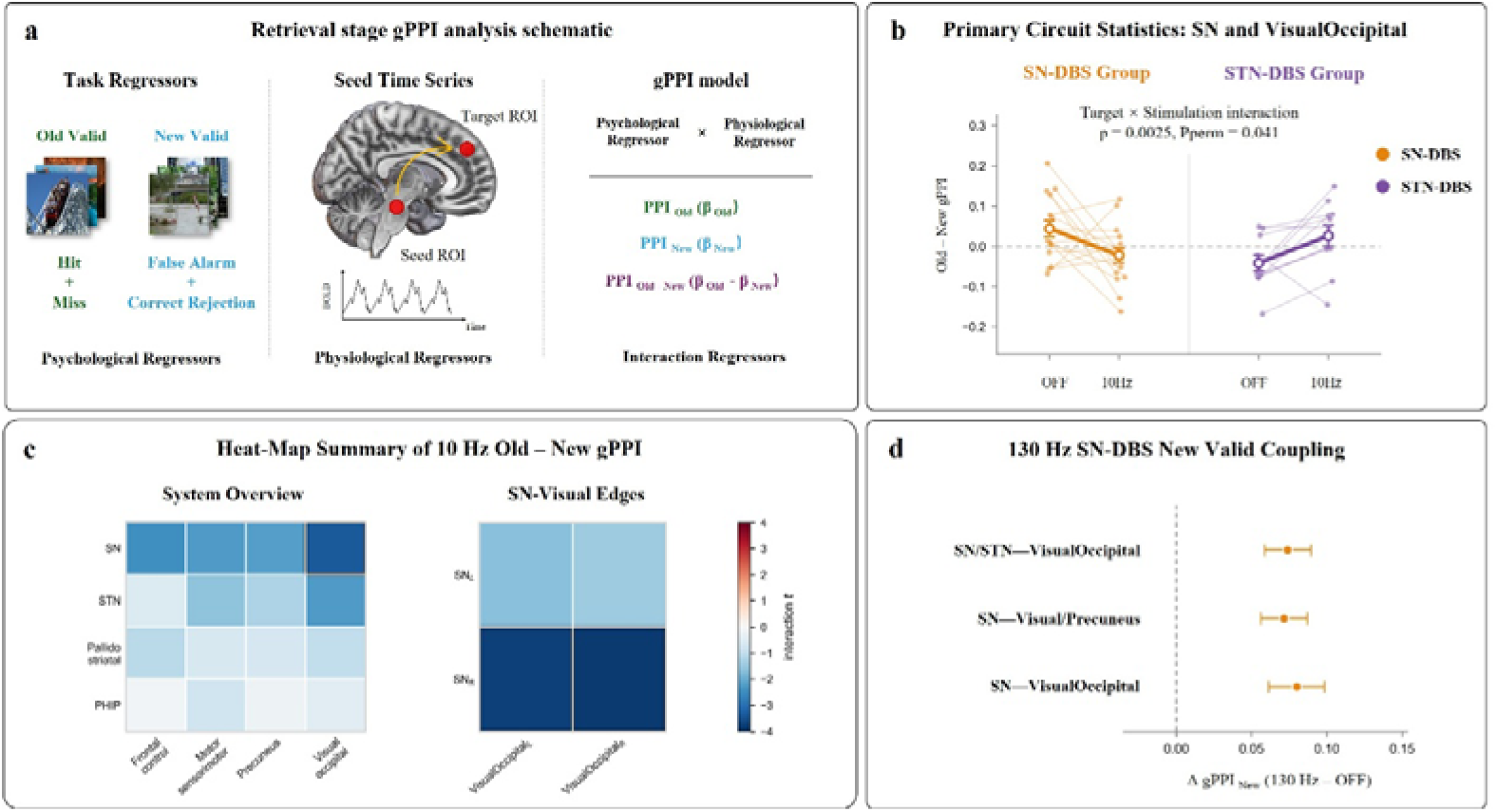
Event-related gPPI identifies target-dependent modulation of SN–visual retrieval circuitry. **(a)** Schematic of the retrieval-stage gPPI analysis. Event regressors were defined for valid old and valid new image presentations during retrieval. Seed ROI time series were extracted from predefined deep-brain and cortical ROIs, and gPPI interaction terms were estimated for new-image coupling and old–new coupling. **(b)** Primary 10 Hz circuit result for SN–VisualOccipital old – new gPPI. Thin lines and small points indicate participant-level values under OFF and 10 Hz stimulation; large points and error bars indicate group-level estimates. SN-DBS decreased, whereas STN-DBS increased, old–new SN–VisualOccipital coupling from OFF to 10 Hz. The target × stimulation interaction survived max-statistic permutation correction. **(c)** Heat-map summary of 10 Hz old–new gPPI target-group interactions. Left, system-level screen across predefined seed and cortical target systems. Right, edge-level localization within the SN–VisualOccipital circuit. Effects were concentrated in SN–VisualOccipital pathways, with the strongest interactions involving right SN coupling with bilateral VisualOccipital targets. Colors indicate interaction t statistics. **(d)** Complementary 130 Hz result in the SN-DBS group. SN-DBS increased new-image gPPI in SN/STN–VisualOccipital, SN–Visual/Precuneus, and SN–VisualOccipital posterior retrieval circuits. Points indicate mean stimulation-induced changes relative to OFF, and horizontal bars indicate confidence intervals. q values are FDR-corrected within the analysis family unless otherwise indicated.

The primary event-related circuit result emerged for SN–VisualOccipital old–new gPPI at 10 Hz. In this predefined circuit, SN-DBS and STN-DBS showed opposite stimulation-induced changes relative to OFF (**Fig.5 b**). SN-DBS reduced old–new SN–VisualOccipital coupling at 10 Hz, whereas STN-DBS increased this coupling. The target × stimulation interaction was significant in the primary circuit analysis (mean ΔSN = −0.067, mean ΔSTN = 0.068, βSN−STN = −0.134, t = −3.36, p = 0.0025, max-stat permutation p = 0.041). Thus, during retrieval, 10 Hz SN-DBS and 10 Hz STN-DBS exerted opposing effects on how SN–visual circuitry differentiated old from new images.

An edge-level follow-up localized this primary effect to the right SN and bilateral visual occipital targets. An edge refers to a specific seed–target ROI pair; this analysis therefore tested which hemispheric SN–visual connections accounted for the system-level SN–VisualOccipital interaction. At 10 Hz, right SN–left VisualOccipital old–new gPPI decreased in the SN-DBS group and increased in the STN-DBS group (mean ΔSN = −0.109, mean ΔSTN = 0.093, β = −0.202, q = 0.004). The same pattern was observed for right SN–right VisualOccipital coupling (mean ΔSN = −0.109, mean ΔSTN = 0.091, β = −0.200, q = 0.004). Because old–new gPPI indexes the relative difference in coupling during old versus novel retrieval items, these edge-level effects indicate that 10 Hz DBS modulated right SN–visual discrimination-related coupling in opposite directions depending on whether stimulation was delivered to SN or STN **(Fig.5 c**).

Target-group interactions for 10 Hz old–new gPPI were concentrated in circuits involving SN and visual occipital targets, with the strongest effects in right SN–VisualOccipital edges (**Fig.5 c**). Pallidostriatal, parahippocampal and frontal-control circuits did not show comparable corrected target-group interactions in the primary analysis. This pattern suggests that event-related modulation of old–new retrieval processing was most prominent in the SN–visual occipital pathway.

A complementary result was observed at 130 Hz for new-image coupling. In the SN-DBS group, 130 Hz stimulation significantly increased new-image gPPI in three posterior retrieval circuits: SN/STN–VisualOccipital (mean Δ = 0.074, t = 4.80, q = 0.010), SN–Visual/Precuneus or posterior retrieval regions (mean Δ = 0.072, t = 4.62, q = 0.010), and SN–VisualOccipital (mean Δ = 0.080, t = 4.30, q = 0.011; **Fig.5 d**). These findings indicate that, at 130 Hz, SN-DBS enhanced coupling between nigral/subthalamic seeds and posterior visual–retrieval systems during processing of new images.

Several control analyses constrained the interpretation of these event-related findings. Analyses of cortical target systems alone, and analyses focused on deep/hippocampal seeds to frontal control targets, did not yield FDR-corrected effects. In addition, direct gPPI false alarm reduction associations did not survive correction. Therefore, the event-related gPPI results should be interpreted as convergent evidence that DBS modulates retrieval-related SN–visual circuitry, rather than as direct evidence that event-related coupling independently predicts false alarm reduction. Together with the resting-state FC and ALFF findings, these results support a model in which SN-DBS engages visual and posterior retrieval systems during new-item processing and old – new discrimination.

### HC event-related fMRI reference supported posterior visual–parietal retrieval systems

As a healthy reference for task-evoked old/new retrieval processing, we applied the same retrieval-stage event-related GLM to the 16 HC participants. At the same whole-brain threshold used for the patient imaging analyses, New > Old produced circumscribed activation in left lateral occipital/occipito-temporal cortex and left precentral/postcentral cortex. In contrast, Old > New produced a broader posterior retrieval pattern involving bilateral precuneus/cuneus and left inferior parietal/angular cortex, together with right precentral/postcentral, supplementary motor and superior frontal regions (***Supplementary Fig. 4***). Thus, healthy participants engaged posterior visual–parietal and sensorimotor-retrieval systems during the same old/new recognition task, supporting the interpretation that the DBS effects observed in patients converged on task-relevant retrieval systems rather than on isolated subcortical nuclei. We next applied the same ROI-to-ROI HRF-deconvolved gPPI framework to the HC task-fMRI data, focusing on the predefined deep-to-cortical circuits corresponding to the patient gPPI analysis. All HC sessions passed retrieval and ROI quality control. However, planned one-sample tests did not identify corrected nonzero HC cortico-subcortical gPPI effects for either new-image coupling or old–new coupling. Therefore, the HC gPPI results were treated as a reference/control analysis rather than as an independent positive connectivity finding. The absence of corrected HC deep-to-cortical task-gPPI effects suggests that the patient SN–visual and posterior retrieval gPPI effects may reflect DBS-induced recruitment or reweighting of a subcortical–posterior cortical retrieval pathway in PD, rather than simple amplification of a robust task-coupling pattern already detectable in HCs.

To further account for inter-individual variability in electrode implantation sites and estimated stimulation fields, we performed an exploratory connectome-based fiber-filtering analysis using Lead-DBS. This analysis related 10 Hz–induced changes in overall recognition accuracy to structurally connected fiber pathways. Positively weighted streamlines, associated with greater accuracy improvement, projected predominantly toward prefrontal regions, whereas negatively weighted streamlines, associated with poorer accuracy change, preferentially targeted sensorimotor and parietal cortices, including postcentral and precentral regions (***Supplementary Fig. 5***). These exploratory findings suggest that individual variability in low-frequency DBS effects may be partly explained by engagement of dissociable cortico-subcortical pathways.

## Discussion

### SN-DBS reduces false recognition, whereas 10 Hz STN-DBS increases false recognition

In this study, we tested whether different subcortical stimulation targets can differentially regulate human recognition memory. The principal finding is that SN-DBS and STN-DBS produced opposing effects on episodic recognition. SN-DBS improved recognition sensitivity at both 10 Hz and 130 Hz, whereas STN-DBS, particularly at 10 Hz, reduced recognition performance. This dissociation was not explained by motor state, response speed or a global shift in response criterion: the behavioral effect survived adjustment for UPDRS-III, reaction-time measures were not significantly modulated, and bias c did not show a corresponding stimulation-dependent pattern. Instead, the effect was expressed most clearly in false alarm rate. SN-DBS reduced false recognition of novel images at both frequencies, whereas 10 Hz STN-DBS increased false recognition.

This behavioral profile is important because it indicates that SN-DBS did not simply amplify memory strength or increase the tendency to respond “old”. If stimulation had produced a nonspecific response bias, one would expect systematic changes in bias c or reaction time. Instead, SN-DBS improved discrimination primarily by reducing the probability that novel but perceptually similar images were misclassified as previously studied. Recognition memory depends on the ability to bind item information to contextual details and to reject familiar-looking lures when they lack the appropriate episodic context. This interpretation is consistent with contextual binding theories of episodic memory and with models in which prefrontal–hippocampal interactions support retrieval monitoring and the selection of appropriate mnemonic evidence^1, 2, 36^.

These findings also refine the broader human memory-stimulation literature. Direct stimulation of memory-related regions has produced both improvement and impairment depending on stimulation site, timing, state and protocol. Entorhinal stimulation during learning was reported to enhance spatial memory, whereas stimulation of entorhinal/hippocampal sites at other parameters impaired memory, and closed-loop temporal cortical stimulation improved recall only when delivered during low-memory states^15–17^. Our results extend this principle to deep PD targets: cognitive DBS effects are not uniform properties of stimulation, but depend on target identity, frequency and the cognitive operation being tested. Here, the most strongly modulated operation was not hit detection but false-recognition suppression, i.e., the accurate detection of novel stimuli that was highly similar to those presented at encoding.

### Imaging mechanisms: an OFF false-recognition circuit and DBS target-dependent visual retrieval modulation

The imaging results suggest that the behavioral dissociation arose from modulation of a pre-existing false-recognition/correct novelty detection circuit rather than from a uniform change in global connectivity. Under OFF, false alarm rate was associated with intrinsic coupling among subcortical, medial temporal, frontal and posterior visual regions, including SN, STN, pallidal, hippocampal/parahippocampal, superior frontal/inferior frontal and occipital/lingual/fusiform nodes. This circuit architecture is biologically plausible. Episodic memory retrieval requires medial temporal representations, frontal monitoring and posterior cortical reinstatement, while basal ganglia–thalamic loops can influence large-scale cortical interactions^1, 2, 10, 11^.

The SN-DBS connectivity results indicate that stimulation-related benefit was expressed as individualized modulation of this OFF false-recognition circuit. Connections that were positively associated with false alarm rate under OFF tended to decrease in participants with greater SN-DBS-related false alarm reduction, whereas connections that were negatively associated with false alarm rate under OFF tended to increase. This directional correspondence supports the interpretation that SN-DBS shifted a vulnerability-state circuit toward a less false-recognition-prone configuration. The parahippocampal–superior frontal gyrus connection was particularly notable: it appeared in the OFF false alarm circuit, was modulated in relation to SN-DBS false alarm reduction at both 10 Hz and 130 Hz and retained a DBS target-dependent effect in baseline-adjusted sensitivity analysis. This pathway links medial temporal mnemonic evidence with frontal recollection processes^35^ and frontal decision-control processes needed to reject similar lures^37^.

A second mechanistic component concerns neurochemically heterogeneous nigral signaling during novelty and familiarity evaluation. A dopamine-related account remains plausible: midbrain SN/VTA activity codes stimulus novelty, putative human dopaminergic SN neurons discriminate novelty from familiarity and predict declarative-memory formation, and VTA/SN dopamine can facilitate associative memory encoding in entorhinal cortex^12, 21, 38^. However, the stimulated region in the present study encompassed SN, and 3T fMRI could not resolve dopaminergic the substantia nigra pars compacta (SNc) from GABAergic the substantia nigra pars reticulata (SNr) contributions. The SNr is a major basal ganglia output nucleus whose GABAergic projection neurons target multiple diencephalic and brainstem structures, including the superior colliculus^39, 40^. This anatomy is relevant to the present findings because false alarms in our task required visually similar lures to be rejected based on mnemonic evidence, and SN-DBS effects converged on posterior visual, parieto-occipital and SN–VisualOccipital retrieval circuits. The superior colliculus is not only an oculomotor structure; it contributes to visual spatial attention and can bidirectionally shape frontal choice activity^41, 42^. We therefore interpret SN-DBS not as a purely dopaminergic manipulation, but as stimulation of a neurochemically heterogeneous nigral node in which dopamine-dependent novelty/familiarity signaling and SNr GABAergic output gating may jointly bias retrieval-stage visual evidence. Under this account, SN-DBS could reduce false recognition by increasing the separation between mnemonic familiarity and visual similarity, while SNr output pathways may regulate which visual evidence is allowed to influence frontal–medial temporal retrieval decisions.

The ALFF and gPPI findings further suggest that this circuit-level effect was expressed through posterior visual and sensorimotor–visual systems. At 10 Hz, greater false alarm reduction under SN-DBS was associated with increased ALFF in dorsal parieto-occipital and postcentral regions, whereas STN-DBS showed the opposite relationship in right occipital visual cortex. Event-related gPPI converged on the same axis: at 10 Hz, SN-DBS and STN-DBS drove opposite changes in old–new SN–VisualOccipital coupling, with edge-level effects localized to right SN and bilateral visual occipital targets. At 130 Hz, SN-DBS increased new-image coupling and posterior visual/precuneus retrieval regions. These results suggest that DBS influenced not only a resting-state vulnerability circuit but also retrieval-stage processing of visual mnemonic evidence^43, 44^. The HC task-fMRI reference further supports this interpretation. Although HCs were not part of the DBS causal model, their old/new recognition activation pattern involved posterior visual–parietal and sensorimotor-retrieval regions, including precuneus/cuneus and inferior parietal/angular cortex. These regions overlap at the systems level with the posterior visual, parieto-occipital and sensorimotor systems implicated by the patient ALFF and task-gPPI findings. Thus, the patient DBS effects should not be interpreted as modulation of an isolated deep nucleus, but as modulation of distributed retrieval systems engaged by the task itself. At the same time, the HC deep-to-cortical gPPI analysis did not reveal corrected nonzero effects in the predefined primary circuits. This negative reference result is informative but should be interpreted cautiously: it does not prove that SN/STN–visual or medial temporal–frontal pathways are absent in healthy recognition memory. Rather, these pathways were not detectably task-modulated in HCs under the present ROI-to-ROI gPPI model and sample size. In this context, the patient SN–visual gPPI effects are best interpreted as DBS-induced recruitment or reweighting of a subcortical–posterior retrieval pathway in PD, rather than simple amplification of a robust HC task-coupling pattern. The recent SCAN framework is relevant here because PD-relevant subcortical targets, including SN, STN and pallidal structures, are strongly linked to somato-cognitive action networks rather than being purely motor nodes^31, 45^. Our findings extend this idea by showing that subcortical DBS targets can influence visual mnemonic decisions through sensorimotor–visual and posterior retrieval systems.

The STN findings should be interpreted differently from the SN findings. Behaviorally, 10 Hz STN-DBS increased false alarms and reduced recognition sensitivity, but the predefined resting-state ΔFC–false alarm reduction analysis did not reveal a corrected STN-specific FC–behavior coupling. Thus, we should not claim that a single STN connectivity pathway caused false recognition. Instead, the convergent ALFF and gPPI results suggest that 10 Hz STN-DBS may perturb retrieval-related visual evidence processing. This interpretation aligns with the known role of the STN in hyper direct frontal control circuits, conflict processing, stopping and cognitive regulation^23^. Importantly, this does not mean that low-frequency STN stimulation is generally harmful for cognition. Prior studies have shown that theta-range STN activity or stimulation can support aspects of cognitive switching and working memory under specific task demands and patient groups^27, 28, 46, 47^. The present result is more specific: in a recognition-memory task requiring rejection of visually similar lures, 10 Hz STN-DBS impaired the system mediating correct identification of novel, similar lures.

The STN result should be interpreted in relation to the medial temporal–frontal false-recognition circuit. Recognition-memory decisions depend on contextual binding, prefrontal–hippocampal interactions and source-monitoring processes, particularly when familiar-looking lures must be rejected in the absence of sufficient contextual mnemonic evidence^43, 44^. Behaviorally, 10 Hz STN-DBS increased false alarms and reduced recognition sensitivity, suggesting impaired rejection of visually similar lures. The strongest adjusted circuit marker was the parahippocampal–frontal connection, which retained a target-dependent association with false alarm reduction after adjustment for baseline false alarm rate at 10 Hz. This suggests that the opposing effects of SN-DBS and STN-DBS may depend on how stimulation interacts with a PHG–frontal circuit supporting mnemonic evidence evaluation and retrieval monitoring. SN-DBS may shift this circuit away from a false-recognition-prone state, whereas 10 Hz STN-DBS may perturb the same control axis, increasing the likelihood that familiar-looking lures are falsely judged as old^1, 2^. The ALFF and gPPI findings provide convergent evidence for altered posterior retrieval dynamics, but they should be considered secondary because direct gPPI–behavior associations did not survive correction. Thus, the STN interpretation remains inferential and is better framed around target-dependent PHG–frontal circuit modulation than around a demonstrated STN–SNr mechanism.

### A working circuit model of target- and frequency-dependent DBS effects on recognition memory

Together, the behavioral and imaging results support a working circuit model in which DBS influences recognition memory through target-, frequency- and state-dependent modulation of subcortical–cortical networks. This model is intended as a mechanistic framework for interpreting the present findings, rather than as evidence for a single linear causal pathway.

The first component involves a baseline false-recognition circuit linking SN, pallidal and subthalamic seeds with medial temporal, frontal and posterior visual regions. Under OFF, false alarm rate was associated with intrinsic coupling within this circuit. During SN-DBS, greater false alarm reduction was associated with stimulation-induced connectivity changes that were oriented opposite to the OFF false-recognition pattern. This effect was most consistently observed for parahippocampal–superior frontal gyrus connectivity, which was implicated at both stimulation frequencies and retained a target-dependent effect in the baseline-adjusted sensitivity analysis at 10 Hz. These findings suggest that SN-DBS-related reduction of false recognition may involve modulation of a pre-existing circuit supporting false familiarity, mnemonic monitoring and visual evidence evaluation.

The second component distinguishes circuit-level inference from neurotransmitter-specific hypotheses. The dopamine hypothesis explains how nigral stimulation could recalibrate novelty/familiarity evidence during retrieval. The GABA hypothesis explains how SN output could gate visual and orienting-related evidence through superior-collicular, thalamic and frontal pathways. These mechanisms are not mutually exclusive. Electrical stimulation can recruit local neurons as well as afferent and efferent axons, and recent mechanistic work indicates that DBS effects depend on target-specific synaptic and axonal recruitment rather than simple excitation or inhibition of a single cell class^49, 50^. We therefore revise the working model to treat the SN as a composite output node. Dopamine-related SNc/VTA signaling and GABAergic SNr output gating are shown as parallel, inferential mechanisms, whereas the data-supported effects are the target-level modulation of false-recognition connectivity, parieto-occipital ALFF and SN–VisualOccipital retrieval coupling.

A third component involves posterior visual and sensorimotor–visual systems. At 10 Hz, SN-DBS-related false alarm reduction was associated with increased ALFF in dorsal parieto-occipital and postcentral regions, whereas STN-DBS showed an opposing relationship in right occipital visual cortex. Task-state gPPI provided convergent evidence: 10 Hz SN-DBS and STN-DBS produced opposite changes in old–new SN–VisualOccipital coupling, and 130 Hz SN-DBS increased new-image coupling in posterior visual/precuneus retrieval circuits. Because direct gPPI–false alarm reduction associations did not survive correction, these task-state findings should be interpreted as convergent evidence that DBS alters retrieval-related visual processing, rather than as an independent behavioral prediction pathway.

This model also clarifies the role of stimulation frequency. The behavioral direction was primarily determined by target: SN-DBS reduced false recognition at both 10 Hz and 130 Hz, whereas 10 Hz STN-DBS increased false recognition. Frequency shaped the imaging expression of these effects. At 10 Hz, target differences were most apparent in ALFF and old–new SN–VisualOccipital gPPI. At 130 Hz, SN-DBS benefit was accompanied by SN-related FC modulation and increased new-image coupling in posterior retrieval circuits. Thus, stimulation frequency should not be interpreted as uniformly beneficial or detrimental for cognition; rather, its effect depends on stimulation target, task demands and baseline circuit state.

In this working model, SN-DBS may reduce false recognition by modulating a baseline false-recognition circuit involving SN, medial temporal, frontal and posterior visual regions. By contrast, 10 Hz STN-DBS may perturb fronto-subthalamic and posterior visual processes required for rejecting visually similar lures. The model remains schematic and hypothesis-generating: it does not establish dopamine release, anatomical directionality or a fully independent predictive biomarker. (**Fig. 6**)

**Fig. 6.**
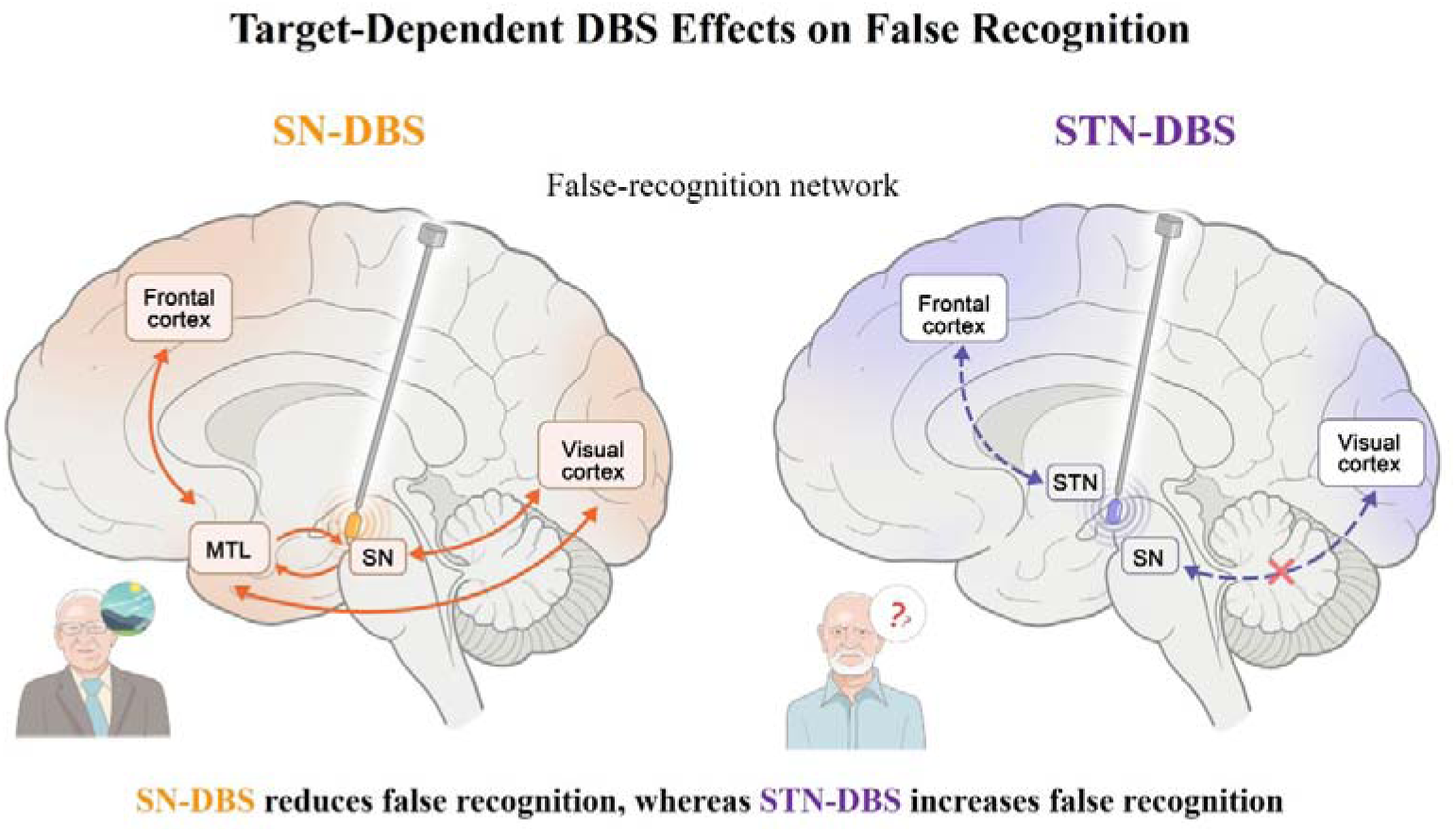
Target-dependent DBS modulation of false-recognition circuits. Schematic working model illustrating how SN-DBS and STN-DBS differentially modulate false recognition during recognition memory. In the SN-DBS condition, stimulation of the substantia nigra engages a false-recognition network involving the SN, medial temporal lobe, frontal cortex and posterior visual cortex. This network-level modulation is proposed to shift patients away from a false-familiarity state, thereby reducing false alarms and improving recognition sensitivity. In contrast, 10 Hz STN-DBS is proposed to perturb a frontal–STN–visual pathway involved in evaluating visual mnemonic evidence. This disruption may bias patients toward erroneous familiarity when judging visually similar novel images, resulting in increased false recognition and reduced recognition sensitivity.

**Fig. 7.**
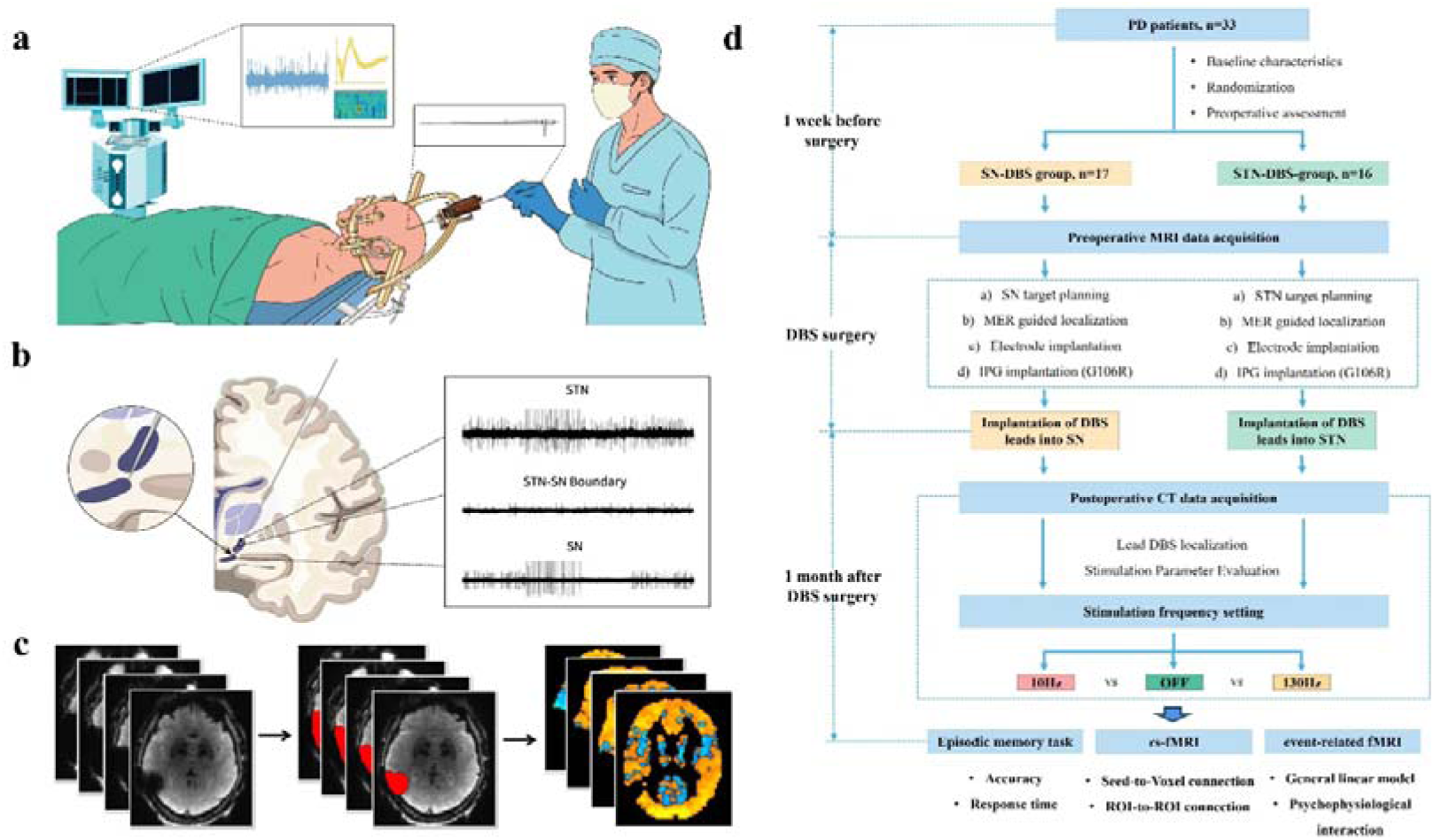
Study protocol. **(a)** Schematic illustration of the DBS surgical procedure; **(b)** Schematic of the DBS lead implantation trajectory with intraoperative MER signals from the ventral STN, dorsal STN, and SN; **(c)** Schematic illustration of DBS electrode artifact masking during fMRI preprocessing; **(d)** Experimental flowchart. A total of 33 patients with PD were enrolled and underwent preoperative data collection, random group assignment, target planning, postoperative evaluation, and fMRI scanning. The entire workflow was jointly overseen and carried out by two experienced neurosurgeons. All procedures were fully explained to participants prior to enrollment, and participants were free to withdraw from the study at any time on a voluntary basis. Participants who withdrew were excluded from all analyses.

Clinically, this distinction is important. Conventional DBS programming in PD is optimized for motor benefit, and high-frequency STN-DBS is a well-established treatment for motor complications^19, 25^. Cognitive impairment is also common in PD, but it is not addressed by the same target–frequency logic that guides motor programming^18^. In the present study, the stimulation condition most strongly associated with motor therapy is not the same as the condition producing cognitive benefit. This supports the idea that motor treatment and cognitive modulation may require different targets^51^, different frequencies and different network pathways. It also suggests that future clinical strategies could consider dual-target or multi-contact programming, in which one set of parameters is optimized for motor symptoms, and another is designed to support cognitive function. Such a strategy would require prospective validation, chronic stimulation testing and safety evaluation, but the present data provides a circuit-level rationale for moving beyond motor-only DBS programming in PD.

Several limitations should be considered. First, all patients had PD, and dopaminergic degeneration may shape both baseline false-recognition vulnerability and stimulation response. The findings therefore cannot be directly generalized to healthy memory systems. Second, the stimulated SN region cannot be cleanly separated into SNc and SNr using 3.0T fMRI, and we did not directly measure dopamine release. Dopamine and novelty provide a strong mechanistic framework, but the present data do not prove a dopaminergic mechanism. Third, the vulnerability score marked SN-DBS response before baseline adjustment, but most composite effects were attenuated after accounting for baseline false alarm rate; only the parahippocampal–superior frontal gyrus connection retained a target-dependent adjusted effect. Fourth, gPPI analyses showed target-dependent event-related circuit modulation, but direct gPPI–false alarm reduction associations did not survive correction. Fifth, DBS-fMRI is technically challenging, and although susceptibility artifacts, motion and invalid task sessions were controlled, residual effects may remain. Finally, the study used acute stimulation conditions; future work should test whether the same cognitive effects persist under chronic stimulation and whether out-of-sample circuit markers can predict individual cognitive response.

In summary, our findings show that DBS can bidirectionally regulate human recognition memory through target-specific engagement of subcortical–cortical circuits. SN-DBS-related false alarm reduction was associated with modulation of a baseline false-recognition circuit involving SN, medial temporal, frontal and posterior visual regions. STN-DBS, particularly at 10 Hz, produced the opposite behavioral effect and was associated with divergent visual retrieval dynamics. These findings identify the SN as a candidate node for pro-cognitive neuromodulation and suggest that the cognitive effects of DBS depend on target, frequency and pre-stimulation circuit state.

## Methods

This was a single-blind, randomized prospective study designed to investigate the differential effects of STN-DBS and SN-DBS on recognition-based episodic memory. The study was conducted in strict adherence to relevant national and international guidelines, including the principles outlined in the Declaration of Helsinki. The study was initiated only following approval by the institutional research ethics board (Ethics Committee of Beijing Tiantan Hospital, Approval No.: KY2024-025-02) and registration at ClinicalTrials.gov (Identifier: JQ23038-2).

Inclusion criteria for PD patients were: (1) clinical diagnosis of idiopathic PD; (2) voluntary participation with written informed consent; (3) age between 50 and 70 years; and (4) normal or corrected-to-normal hearing and vision, with sufficient cognitive ability to complete all experimental procedures. Exclusion criteria included: (1) severe psychiatric, cognitive, or psychological disorders; (2) contraindications to neurosurgery; or (3) contraindications to magnetic resonance imaging (MRI) scanning. The 16 HC participants were matched for age range, confirmed free of neurological disorders via screening, and had normal cognitive function (***Supplementary Materials Table 1***).

Baseline demographic and clinical characteristics were collected for all enrolled PD patients, including age, gender, disease duration, Hoehn and Yahr stage, and levodopa equivalent daily dose (LEDD). Prior to the experiment, all participants completed a series of standardized assessments: UPDRS-III was administered in the medication-on state; MoCA and Mini-Mental State Examination (MMSE) were used to evaluate global cognitive function; and the 14-item Hamilton Anxiety Rating Scale (HAMA) and 24-item Hamilton Depression Rating Scale (HAMD) were employed to assess anxiety and depression levels, respectively. All evaluations were performed by two neurosurgeons with specialized training in neuropsychological assessment.

Following completion of baseline clinical and neuropsychological assessments, the PD patients were randomly assigned in a 1:1 ratio to one of two surgical groups. In the SN-DBS group, the deepest contact of the implanted electrode was targeted at the SN. In the STN-DBS group, the deepest contact was positioned within the ventral subdivision of the STN. Randomization was conducted using computer-generated allocation surgical procedures.

### DBS surgery

All patients underwent standard frame-based stereotactic DBS implantation surgery (**Fig.8 a**). Surgical procedures were performed as previously described^52, 53^. Briefly, awake bilateral STN-DBS surgery was conducted using standard functional stereotactic techniques combined with microelectrode recording. For surgical planning, preoperative CT scans acquired with a stereotactic frame were co-registered with preoperative MRI data in the planning system (Surgiplan software, ELEKTA Instruments AB, Switzerland). The STN targeting method followed protocols previously reported in the literature^52, 54^. The STN was visualized on T2-weighted MRI sequences, while the surgical trajectory was planned using T1-weighted sequences with gadolinium enhancement. During microelectrode recording (sampling rate: 44 kHz), a single microelectrode was positioned at the center of the Ben-Gun arrangement integrated into the microelectrode recording system (Neuro Omega, Alpha Omega Engineering, Israel). Recordings were initiated 10 mm above the target site and advanced incrementally in 0.3–0.5 mm steps. Entry into the STN was characterized by two key electrophysiological features: a marked increase in background neural activity and distinct single-unit firing patterns. After exiting the STN, the microelectrode was advanced further into the SN to define the boundary between these two structures. Identification of SN entry was performed via visual assessment by two electrophysiological specialists. Typically, the SN is distinguished by relatively lower-amplitude signals accompanied by a more regular spiking pattern^54^. Furthermore, high-frequency, low-amplitude microstimulation was found to suppress single-unit firing within the SN, a phenomenon that facilitated accurate localization of the SN region^56, 57^(**Fig.8 b**).

Two quadripolar DBS electrodes (Model L301C or L302C; Beijing PINS Medical Co., Ltd., China/Model B33005 or B33015, Medtronic, Minneapolis, MN, USA) were bilaterally implanted into the STN under microelectrode recording guidance. Models L301C/Model B33005 and L302C/Model B33015 represent the short and long electrode versions, respectively. The choice between the two was determined preoperatively based on the patient’s clinical presentation: specifically, patients with dystonia, gait disturbance, or other symptoms suggesting that the stimulation field may need to extend into the zona incerta or adjacent structures underwent implantation of the long electrodes (L302C or B33015) to ensure adequate coverage of the zona incerta. In contrast, the short electrodes (L301C or B33005) were chosen when no such extended coverage was required (***Supplementary Materials* *Figure 1***). For patients in the STN group, the tip of the first DBS contact was placed exactly at the depth of the STN lower boundary; for patients in the SN group, this tip was positioned 2.5–3.0 mm below the SN upper boundary. Intraoperative test stimulation was delivered to evaluate symptom improvement and adverse events in all patients. A 3T MRI-compatible neurostimulator (G106R, Beijing PINS Medical Co., Ltd., China or B35300 Percept RC; Medtronic, Minneapolis, MN, USA) was connected to the DBS leads via extension cables (Model E202C, Beijing PINS Medical Co., Ltd., China or Model 37086; Medtronic, Minneapolis, MN, USA) and subsequently implanted into the subclavian fossa after electrode implantation. According to the manufacturer’s specifications, these systems are MRI-conditional at 3.0 T, permitting full-body MRI scans without the need for device deactivation or parameter adjustments prior to scanning (https://en.pinsmedical.com).

### DBS leads localization

Approximately one month after surgery, a CT scan (0.625 mm slice thickness, GE Medical, USA) was performed. Electrode localization was conducted using Lead-DBS v3.2^29^ (https://www.lead-dbs.org), a toolbox based on MATLAB (R2022b, MathWorks, Natick, MA, USA), following established procedures. Specifically, postoperative CT images were linearly co-registered to presurgical structural MRI using SPM12 (Wellcome Department of Imaging Neuroscience, University College London, U.K.). Presurgical MRI scans were then normalized to Montreal Neurological Institute (MNI) space using Advanced Normalization Tools. Electrode trajectories were automatically reconstructed using the PaCER^58^ algorithms and manually refined when necessary.

### Stimulation parameter

All stimulation parameters were determined by a board-certified neurosurgeon specializing in functional neurosurgery. Three stimulation conditions were tested: 10 Hz stimulation, 130 Hz stimulation, and the OFF A monopolar configuration was adopted, with active lead contact as the cathode and the neurostimulator as the anode. In both groups, the deepest electrode contact was used for stimulation. The stimulation current intensity for each condition was individually titrated to the highest amplitude that did not induce adverse effects under both low- and high-frequency stimulation, while the pulse width was kept constant at 60 μs. The current amplitude was identical between the two stimulation frequencies to maintain a constant of the volume of tissue activated. In patients implanted with Medtronic leads, stimulation during 3T MRI was delivered in a bipolar configuration, because the MRI-compatible mode for this system did not allow monopolar stimulation under the scan conditions used here. Contact selection in these patients was therefore constrained to bipolar stimulation. To reduce variability introduced by differences in stimulation geometry, electrode placement and stimulation parameters were chosen with particular attention to the estimated volume of tissue activated, with the aim of keeping stimulation as confined as possible to the intended anatomical target and limiting spread into adjacent structures. The order of the three stimulation conditions was randomized across participants to minimize potential order effects. Lead-DBS v3.2 was used to calculate the individual volume of tissue activated for each patient under the predefined stimulation parameters. Based on these simulations, the estimated volume of tissue activated was used to determine the appropriate range of stimulation current for each patient. For the STN-DBS group, stimulation parameters were adjusted such that the volume of tissue activated did not extend beyond the caudal (lower) boundary of the STN. Conversely, for the SN-DBS group, stimulation intensity was titrated to ensure that the volume of tissue activated remained strictly within the SN and did not extend above its rostral boundary (***Supplementary Materials* *Figure 2***).

### Episodic memory task design

The episodic memory task performed during event-related fMRI was adapted from the designs of Hung-Yu Chen et al.^59^ and Emily A Mankin et al.^60^ and implemented in E-Prime 3.0 (Psychology Software Tools, Pittsburgh, PA, USA). Specifically, the task consisted of three phases: encoding, interference, and retrieval. During the encoding phase, 36 pictures were presented in a randomized order; each trial began with a 0.5-s fixation cross, followed by a 1-s picture presentation and a 0.5-s blank screen. Participants were repeatedly instructed to encode as many perceptual details of each picture as possible. In the interference phase, 20 random digits were presented sequentially at the center of the screen, and subjects performed a parity judgment (odd/even) via key press for each digit; this phase lasted 60 s, with the same fixation and blank-screen timing as in the encoding phase. During the retrieval phase, 72 pictures were shown in randomized order: the 36 previously encoded images and 36 foils that were perceptually similar but differed in specific details. Participants were asked to discriminate old from new based on the encoded details and to respond within 2500 ms. Inter-stimulus intervals (ISI) were jittered between 2,000 and 5,000 ms (mean = 3500 ms). A total of 216 unique images were used, all drawn from the Open Affective Standardized Image Set (OASIS) dataset with normative ratings for arousal^61^, (http://www.benedekkurdi.com/#oasis). The stimulus sets used across the three DBS conditions were non-overlapping; each set was matched for mean valence and arousal to ensure equivalent psychological properties across conditions.

### Task performance assessment

All behavioral testing was conducted under single-blind conditions, with participants unaware of the stimulation setting applied in each session. Retrieval responses were classified according to standard old/new recognition outcomes. Previously encoded images judged as old were coded as hits, whereas previously encoded images judged as new were coded as misses. Novel foil images judged as old were coded as false alarms, whereas novel foil images judged as new were coded as correct rejections.

Trials with no response or response times shorter than 200 ms were excluded from signal-detection analyses. For each participant and stimulation condition, we calculated overall recognition accuracy, hit rate, false alarm rate, correct rejection rate, Pr, d′ and response bias. Hit rate was defined as hits divided by the number of valid old-image trials, and false alarm rate was defined as false alarms divided by the number of valid new-image trials. Recognition sensitivity was quantified as d′ = z(hit rate) − z(false alarm rate). Pr was calculated as hit rate − false alarm rate. Response bias c was calculated from the z- transformed hit and false alarm rates. Extreme hit or false alarm rates of 0 or 1 were corrected before z transformation using a standard log-linear correction. Response-time analyses were performed for correct trials only and were calculated separately for hits and correct rejections. Sessions with overall recognition accuracy below 30% were excluded because performance at this level was considered insufficient for reliable interpretation of recognition behavior.

False alarm reduction was defined as false alarm rate under OFF minus false alarm rate under active stimulation, such that positive values indicated reduced false recognition during active DBS. This measure was used in subsequent brain–behavior analyses linking stimulation-induced imaging changes to behavioral improvement.

### Acquisition of fMRI data

All PD patients underwent one T1-weighted structural MRI scan and three separate functional MRI sessions in the medication-off state, one for each DBS stimulation condition (OFF, 10 Hz, and 130 Hz). Each functional session included a resting-state fMRI scan and an event-related fMRI scan (7 min 36 s per run). In contrast, HCs completed a single session consisting of only a T1-weighted structural scan and an event-related fMRI scan. Structural and fMRI data were acquired on a 3T Siemens Prisma scanner (Siemens Healthcare, Erlangen, Germany) using a 64-channel head coil. Visual stimuli were presented via an LCD projector (SA-9900 fMRI Stimulation System, Shenzhen Sinorad Medical Electronics, Inc., Shenzhen, Guangdong, China) and viewed through a mirror attached to the head coil. Structural images were obtained using a 3D T1-weighted MPRAGE sequence (192 sagittal slices, TR = 1560 ms, TE = 1.65 ms, TI = 778 ms, flip angle = 8°, matrix = 256 × 232, slice thickness = 1 mm, voxel size = 1 × 1 × 1 mm). BOLD images were acquired using an echo-planar imaging (EPI) sequence (TR = 2000 ms, TE = 30 ms, flip angle = 90°, FOV = 188 × 188 mm², matrix = 94 × 94, multiband factor = 2, 60 slices, voxel size = 2 × 2 × 2 mm, 222 volumes per run).

### Preprocessing of fMRI data

Magnetic susceptibility artifacts induced by DBS electrodes predominantly affected the left hemisphere, including the inferior parietal, temporal, and occipital lobes, as well as the cerebellum. Preprocessing was performed using SPM12 (Wellcome Department of Imaging Neuroscience, University College London, UK) and MATLAB (R2022b, MathWorks, Natick, MA, USA). Following an initial visual inspection for gross artifacts, functional images were first realigned to correct head motion. Subsequently, manually defined artifact masks were applied to exclude DBS-related susceptibility artifact regions from further preprocessing (**Fig.8 c**), including normalization. The remaining preprocessing steps comprised slice-timing correction, co-registration to each participant’s T1-weighted structural image, normalization to MNI space, and spatial smoothing with a 6-mm full-width at half-maximum Gaussian kernel. Participants exhibiting excessive head motion (>2 mm translation or >2° rotation in any direction) were excluded from further analyses.

### Analysis of fMRI data

Resting-state fMRI preprocessing, denoising and functional connectivity analyses were performed using the CONN toolbox^62^ (version 22a; RRID:SCR_009550) implemented in MATLAB and SPM12. Analyses were performed on preprocessed and denoised BOLD time series after exclusion of participants or sessions failing image-quality or head-motion criteria. Unless otherwise specified, functional connectivity values were Fisher z-transformed before group-level statistical analysis. A priori regions of interest were selected to cover disease-relevant basal ganglia structures, DBS targets and memory-related medial temporal structures. The subcortical seed set included bilateral substantia nigra, subthalamic nucleus, globus pallidus internus, globus pallidus externus, putamen, nucleus accumbens and thalamic/VIM-related regions. Deep nuclei were defined primarily using HybraPD^32^ and DISTAL atlas^33^ masks. Medial temporal regions, including the hippocampus and parahippocampal cortex, and cortical target regions were defined using AAL-90 anatomical labels^34^ where appropriate.

For the baseline resting-state analysis, seed-to-voxel functional connectivity was estimated under OFF. For each priori seed, individual seed time series were correlated with the time series of all brain voxels, and the resulting correlation maps were Fisher-z transformed. Group-level models then tested whether OFF connectivity was associated with recognition sensitivity, indexed by d′, or false alarm rate. These analyses were used to identify baseline connectivity patterns associated with recognition performance and false recognition.

To test whether stimulation modulated the OFF false-recognition circuit, we extracted Fisher z connectivity values from each significant OFF false-alarm-related seed–cluster pair under OFF, 10 Hz and 130 Hz. Stimulation-induced connectivity change was defined as ΔFC = FC (DBS-active) – FC (OFF). Behavioral improvement was indexed by false alarm reduction, defined as false alarm rate (OFF) − false alarm rate (DBS-active), such that positive values indicated reduced false recognition during active DBS. For each stimulation frequency, we tested whether ΔFC was associated with false alarm reduction within each DBS target group and whether these brain–behavior slopes differed between the SN-DBS and STN-DBS groups. These analyses were restricted to the predefined OFF false-recognition connections and were corrected across the planned connection family.

To assess whether the baseline configuration of the OFF false-recognition circuit was associated with subsequent DBS response, we computed a sign-oriented baseline false-recognition circuit-alignment score. This approach was conceptually motivated by connectome-based pattern-scoring methods, in which behavior or disease-related functional connectivity patterns are aggregated into participant-level scores^63–66^. For each participant, OFF Fisher-z connectivity values were extracted from the OFF false alarm-related seed–cluster pairs. Connections positively associated with false alarm rate under OFF were entered with a positive sign, whereas connections negatively associated with false alarm rate under OFF were sign-reversed. The resulting oriented connectivity values were z-scored and averaged across connections. Higher scores therefore indicated stronger alignment with the OFF-connectivity pattern associated with higher false alarm rate. This score was related to false alarm reduction at 10 Hz and 130 Hz, and target differences in alignment–response slopes were tested. Because false alarm reduction was defined as false alarm rate under OFF minus false alarm rate under active DBS, baseline-adjusted sensitivity analyses were performed to determine whether any circuit marker retained target-dependent effects after accounting for baseline false alarm rate.

To assess stimulation-related changes in spontaneous local activity, we performed whole-brain analyses of the amplitude of low-frequency fluctuation (ALFF) using resting-state fMRI data. ALFF maps were computed for each participant and stimulation condition from the preprocessed resting-state BOLD time series within the low-frequency band used for the resting-state analysis. The resulting ALFF maps were entered into group-level models after spatial normalization and smoothing using the same preprocessing pipeline as the functional connectivity analyses. For each active stimulation condition, stimulation-induced ALFF change was defined as ΔALFF = ALFF (DBS-active) – ALFF (OFF). The primary ALFF analysis tested whether ΔALFF was associated with false alarm reduction and whether this ALFF–behavior relationship differed between the SN-DBS and STN-DBS groups. Analyses were performed separately for 10 Hz and 130 Hz stimulation. Whole-brain statistical maps were used to identify regions showing group-specific ALFF–behavior slopes or target-group differences in these slopes. Cluster-level regions identified from the whole-brain analysis were then used for ROI-level extraction to visualize effect sizes and estimate slopes; these ROI extractions were used for descriptive visualization rather than independent statistical confirmation. To contextualize representative ALFF clusters within broader functional networks, additional seed-to-voxel connectivity maps were generated using selected ALFF clusters as seeds under OFF. These connectivity profiles were used descriptively to characterize the intrinsic network context of ALFF regions associated with false alarm reduction.

For event-related fMRI, first-level event-related GLMs were constructed separately for each participant and stimulation condition. Retrieval events were time-locked to image onset and modeled separately for valid old-image and valid new-image trials. Trials with missing responses or invalid behavioral responses were excluded from the event-related connectivity analysis. Six head-motion parameters were included as nuisance regressors, and a 128-s high-pass filter was applied. Event-related connectivity during retrieval was examined using generalized psychophysiological interaction (gPPI) analysis. Retrieval-stage sessions were excluded from gPPI analysis if they had excessive missing responses or insufficient valid old or new retrieval events.Seed and target ROIs were defined as a priori to span deep-brain, medial temporal and cortical systems. The subcortical and medial temporal ROI set included bilateral globus pallidus externus, putamen, globus pallidus internus, subthalamic nucleus, nucleus accumbens, substantia nigra, hippocampus, parahippocampal cortex and amygdala. Cortical ROIs were grouped from AAL-90 anatomical labels into functional systems including dorsolateral prefrontal, inferior frontal, orbitofrontal, medial frontal, motor/sensorimotor, insular, cingulate, parietal, precuneus, visual occipital, fusiform and temporal systems.

For each seed ROI, the extracted BOLD time series was used as the physiological regressor. Psychological regressors modeled valid old-image and valid new-image retrieval events, and interaction terms were estimated for each condition. Two event-related connectivity measures were carried forward: new-image coupling, reflecting connectivity during presentation of novel retrieval items, and old–new coupling, reflecting differential connectivity during old versus new image processing. Stimulation-induced gPPI changes were calculated relative to OFF. Because the resting-state analyses identified PHG–frontal connectivity as the strongest adjusted circuit marker of target-dependent false-alarm modulation, event-related gPPI was treated as a complementary task-state analysis rather than as the primary mechanistic test. Within this framework, we examined whether DBS also altered retrieval-related old–new coupling in predefined subcortical–cortical circuits, including SN–VisualOccipital coupling at 10 Hz. Edge-level follow-up analyses were then used to localize significant circuit effects to specific hemispheric seed–target pairs. A broader circuit screen was performed across the predefined ROI systems to assess whether corrected target-group interactions were concentrated in SN–visual pathways or were also present in pallidostriatal, medial temporal or frontal-control circuits. Complementary analyses tested within-group SN-DBS effects on new-image coupling at 130 Hz. Additional control analyses examined cortical target systems alone, deep/hippocampal seeds to frontal-control targets, and direct associations between gPPI changes and false alarm reduction.

For HCs, the same GLM was applied to the single event-related fMRI session. Valid old-image and valid new-image retrieval events were modeled separately, and invalid retrieval trials were modeled as nuisance events. Six head-motion parameters were included as nuisance regressors, and a 128-s high-pass filter was applied. The primary HC contrasts were New > Old and Old > New. Second-level HC maps were assessed using one-sample t tests and thresholded using the same whole-brain criterion as the patient imaging analyses: voxel-wise p < 0.001 with cluster-level FWE correction at p < 0.05. To determine whether the cortico-subcortical coupling patterns observed in patients were detectable in healthy participants under the same task context, we applied the same ROI-to-ROI HRF-deconvolved gPPI framework to HC task-fMRI data. Valid old-image and valid new-image retrieval events were used as psychological regressors, and new-image coupling and old–new coupling were carried forward as the primary gPPI measures. The HC analysis focused on the predefined deep-to-cortical circuits corresponding to the patient gPPI analysis, including SN/STN–visual occipital, SN/STN–posterior retrieval and hippocampal-system–frontal-control circuits. Because HCs had no stimulation manipulation, HC gPPI effects were tested using one-sample statistics and were interpreted as reference/control estimates rather than stimulation-induced changes. Planned HC gPPI tests were corrected across the predefined circuit family using FDR and max-statistic permutation correction.

### Structural connectivity and fiber filtering

Preoperative T1-weighted MRI and postoperative CT were used to localize DBS leads and reconstruct stimulation fields in Lead-DBS. Electric fields were estimated using the Stimulation Volumes model, and voxels with E-field magnitude below 200 V/m were excluded. Structural connectivity was assessed using the PD-matched normative connectome ppmi_85_ewert_2017. Streamlines were considered connected when they intersected stimulation fields in at least 20% of the E-field volumes. To relate structural pathway engagement to behavior, we quantified each participant’s change in overall recognition accuracy at 10 Hz relative to OFF and performed Lead-DBS fiber filtering using Spearman correlation, with hemispheres mirrored (“mirror sides”) to reduce lateralization-related variance. Significant fibers were identified at an uncorrected α level of 0.05, and streamline weights were visualized as positive (red) or negative (blue) effects. Robustness of the association was evaluated using permutation-based testing with 1,000 permutations (Leave-Nothing-Out permutation strategy), reporting both Spearman and Pearson correlation coefficients. (***Supplementary Materials* *Figure 3***).

### Statistical analysis

Behavioral and clinical statistical analyses were performed in Python 3.12. Motor outcomes were analyzed separately from memory outcomes. UPDRS-III scores were compared across stimulation conditions within each DBS target group using repeated-measures analyses followed by paired comparisons between active stimulation and OFF. Recognition-memory outcomes were analyzed using linear mixed-effects models to account for repeated measurements across stimulation conditions. The primary behavioral outcome was d’ . Secondary outcomes included Pr, hit rate, false alarm rate, response bias c, hit response time, correct-rejection response time and no-response rate. For each outcome, the fixed effects included stimulation target, stimulation frequency and their interaction. Subject identity was included as a random intercept. To control stimulation-dependent motor-state differences, UPDRS-III was decomposed into a between-subject component, defined as each participant’s mean UPDRS-III score across sessions, and a within-subject component, defined as the deviation of each session’s UPDRS-III score from that participant’s meaning. UPDRS-III components were included as covariates. Contrasts compared 10 Hz and 130 Hz stimulation with OFF within each target group and tested target differences in stimulation-induced change. p values from behavioral contrasts were corrected using the Benjamini–Hochberg false discovery rate procedure. Model-estimated effects are reported as β coefficients or percentage-point changes, with 95% confidence intervals. For visualization, subject-level change scores were calculated relative to each participant’s OFF value.

For whole-brain resting-state seed-to-voxel and ALFF analyses, second-level inference was performed within the general linear model framework. Unless otherwise stated, whole-brain imaging results were thresholded at voxel-wise P < 0.001 with cluster-level family-wise error correction at p < 0.05. Extracted connectivity, ALFF and gPPI values were analyzed using linear models testing group-specific behavioral slopes and target-group differences in slopes. For planned extracted-metric analyses, P values were corrected using the Benjamini–Hochberg false discovery rate procedure within the corresponding analysis family. For gPPI circuit analyses, stimulation-induced changes were tested at the predefined circuit level and, where appropriate, followed by edge-level localization analyses. The primary SN–VisualOccipital old–new gPPI interaction at 10 Hz was evaluated using max-statistic permutation correction across the predefined circuit family. Secondary edge-level and broader circuit-screen analyses were corrected using false discovery rate correction within the relevant planned family. Model effects are reported as β coefficients, t statistics, percentage or Fisher z changes where appropriate, together with 95% confidence intervals or corrected p/q values.

## Data Availability

Data is available with reasonable requests to the corresponding authors after the approval of the local IRB.

## Code Availability

All code is available with reasonable requests to the corresponding authors.

## Acknowledgments

This work was supported by the National Natural Science Foundation of China (No. 82471490) and Beijing Natural Science Foundation (No. JQ23038). We also would like to thank Sining Xie, Yuanyuan Jiao, Xiaoya Hu from Beijing Tiantan Hospital and other people who consumed time and energy in the study.

