## Supplementary material for "Substantia Nigra and Subthalamic Nucleus Deep Brain Stimulation Exert Opposing Effects on Novelty Recognition in Parkinson’s Disease"

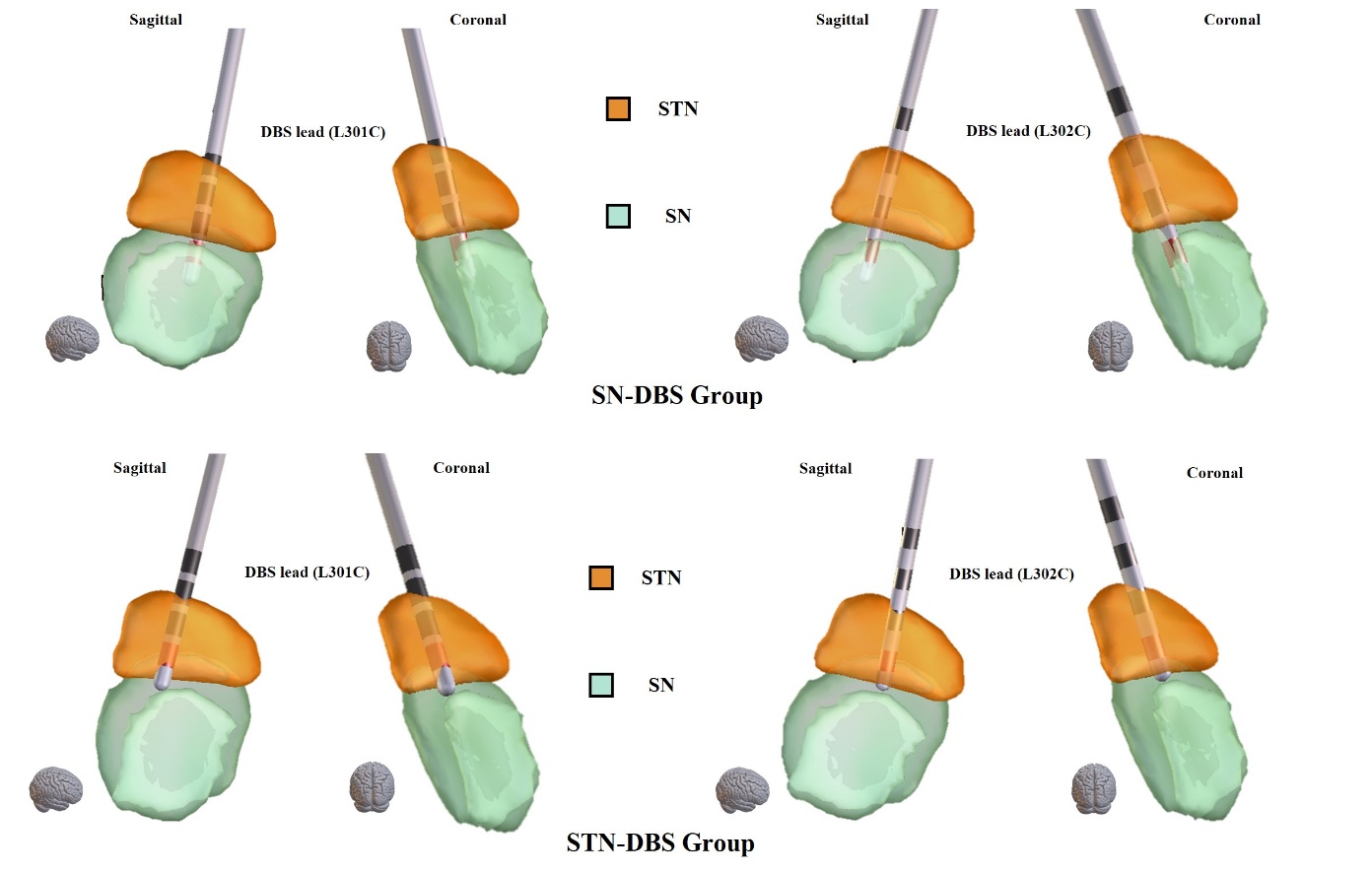

**Supplementary Figure 1 | Representative examples of DBS lead implantation sites.** This panel illustrates the implantation sites using Model L301C and L302C leads (Beijing PINS Medical Co., Ltd., China) as representative examples. All illustrations are derived from participants enrolled in the present study and depict the implantation of the left DBS lead from two different viewing perspectives. The subthalamic nucleus (STN) is shown in orange, and the substantia nigra (SN), encompassing the pars compacta and pars reticulata, is shown in green.

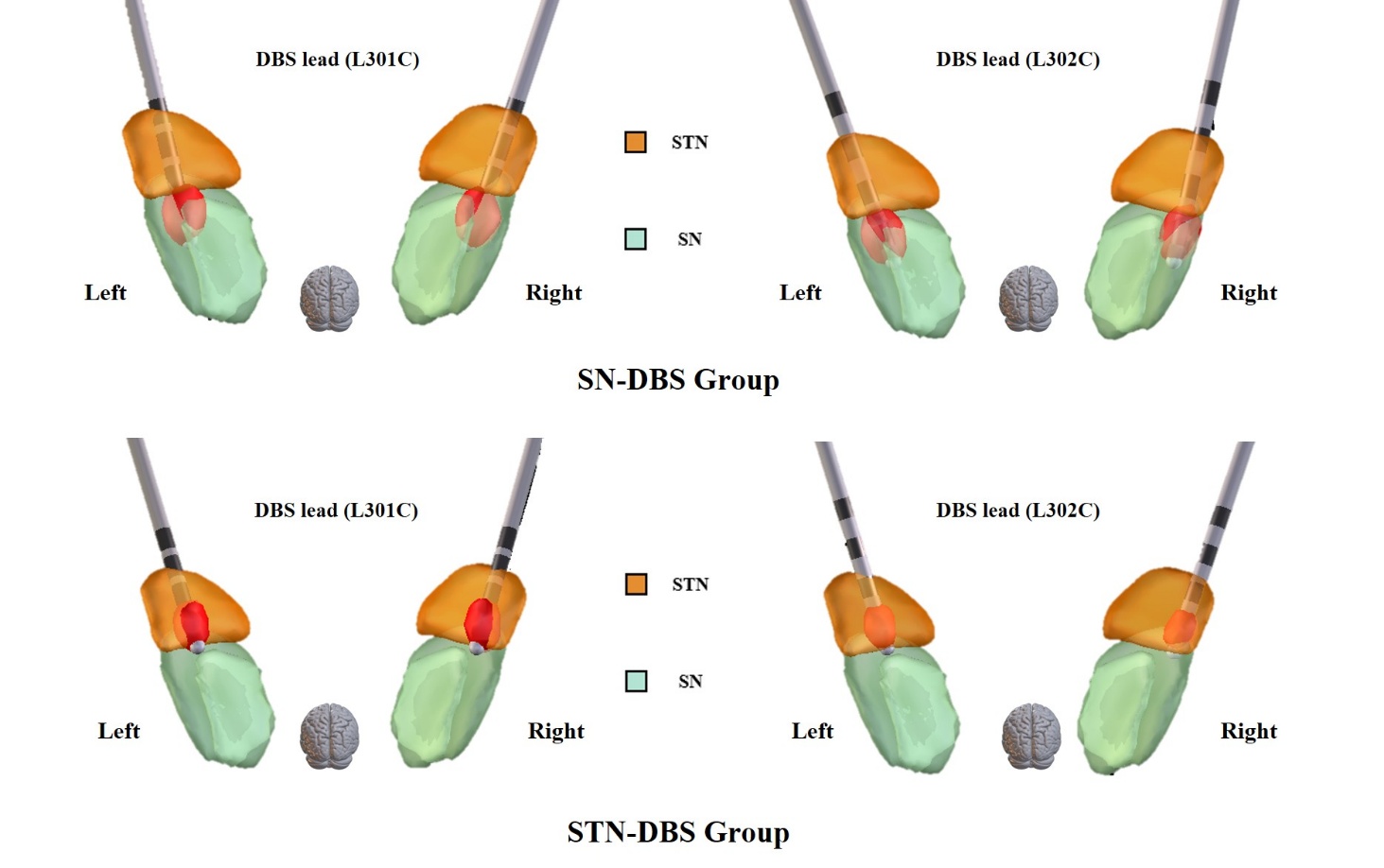

**Supplementary Figure 2 | Schematic illustration of VTA delineation.** This panel illustrates the VTAs corresponding to the stimulation parameters used in the present study for two DBS leads (L301C and L302C) in the SN-DBS and STN-DBS groups. In this panel, STN is shown in orange and SN, which includes the pars compacta and the pars reticulata are shown in green, the volume of tissue activated (VTA) is shown in red.

**Supplementary Table 1 | Baseline Demographic and Clinical Data.** Continuous variables are reported as the mean ± standard deviation (SD); LEED: levodopa equivalent daily dose; UPDRS-III: the Unified Parkinson’s Disease Rating Scale Part III; MoCA: the Montreal Cognitive Assessment; MMSE: the Mini-Mental State Examination; HAMA: the 14-item Hamilton Anxiety Rating Scale; HAMD: the 24-item Hamilton Depression Rating Scale.

| **Characteristics** | **SN-DBS** | **STN-DBS** | **HC** | **p-value (ANOVA)** | **p-value (Within subjects)** | | | |
| --- | --- | --- | --- | --- | --- | --- | --- | --- |
|  |  |  |  |  | **SN-DBS vs STN-DBS** | | **SN-DBS vs HC** | **STN-DBS vs HC** |
| **Age (years)** | 61.44 ± 6.90 | 60.06 ± 8.03 | 58.88 ± 5.58 | 0.580 | 0.846 | 0.591 | | 0.846 |
| **Gender (M/F)** | 9/7 | 14/2 | 11/5 | NA | NA | | NA | NA |
| **Education (years)** | 12.25 ± 5.53 | 12.31 ± 4.80 | 14.19 ± 4.61 | 0.466 | 0.973 | 0.609 | | 0.609 |
| **Disease duration (years)** | 9.56 ± 3.37 | 8.94 ± 2.79 | NA | NA | 0.572 | NA | | NA |
| **Hoehn-Yahr stage (Med-OFF)** | 2.84 ± 0.70 | 2.91 ± 0.64 | NA | NA | 0.794 | NA | | NA |
| **Hoehn-Yahr stage (Med-ON)** | 2.38 ± 0.56 | 2.34 ± 0.44 | NA | NA | 0.862 | NA | | NA |
| **LEED (mg)** | 944.27 ± 341.64 | 962.57 ± 428.96 | NA | NA | 0.898 | NA | | NA |
| **MMSE** | 25.40 ± 4.47 | 26.47 ± 2.50 | 29.06 ± 0.93 | 0.004* | 0.429 | 0.014* | | 0.004* |
| **MoCa** | 22.73 ± 6.85 | 24.00 ± 4.80 | 28.69 ± 1.30 | 0.003* | 0.563 | 0.010* | | 0.006* |
| **HAMA** | 18.27 ± 11.79 | 16.20 ± 10.22 | 7.25 ± 3.80 | 0.004* | 0.612 | 0.009* | | 0.010* |
| **HAMD** | 19.27 ± 14.35 | 11.20 ± 7.36 | 9.69 ± 4.64 | 0.018* | 0.128 | 0.072 | | 0.504 |

**Supplementary Table 2 | DBS electrode information and stimulation parameters for the SN-DBS group.** For “Implantation Depth”, the coordinate origin was defined at the midpoint of the anterior commissure–posterior commissure (AC–PC) line; depths above this reference plan were assigned positive values, whereas depths below were assigned negative values.

| **Number** | **DBS electrodes** | **Neurostimulator** | **Implantation Depth (mm)** | | | **Anode/Cathode** | **Amplitude (mA)** | | **Pulse width (μs)** | |
| --- | --- | --- | --- | --- | --- | --- | --- | --- | --- | --- |
|  |  |  | **Right** | | **Left** |  | **Right** | **Left** | **Right** | **Left** |
| 1 | L302C | G106R | -4.0 | -4.0 | | C+1-/C+5- | 1.7 | 1.7 | 60 | 60 |
| 2 | L302C | G106R | -4.1 | -4.0 | | C+1-/C+5- | 1.5 | 1.5 | 60 | 60 |
| 3 | L301C | G106R | -3.0 | -3.0 | | C+1-/C+5- | 2.2 | 2.3 | 60 | 60 |
| 4 | L302C | G106R | -3.9 | -3.7 | | C+1-/C+5- | 2.2 | 2.0 | 60 | 60 |
| 5 | L301C | G106R | -3.1 | -3.0 | | C+1-/C+5- | 1.8 | 1.8 | 60 | 60 |
| 6 | L302C | G106R | -3.8 | -3.8 | | C+1-/C+5- | 2.5 | 2.4 | 60 | 60 |
| 7 | L301C | G106R | -3.0 | -3.0 | | C+1-/C+5- | 1.3 | 1.3 | 60 | 60 |
| 8 | L302C | G106R | -3.6 | -3.5 | | C+1-/C+5- | 1.8 | 1.8 | 60 | 60 |
| 9 | L302C | G106R | -4.0 | -4.0 | | C+1-/C+5- | 2.1 | 2.2 | 60 | 60 |
| 10 | L302C | G106R | -3.7 | -3.9 | | C+1-/C+5- | 2.0 | 2.0 | 60 | 60 |
| 11 | L301C | G106R | -3.0 | -3.0 | | C+1-/C+5- | 1.5 | 2.0 | 60 | 60 |
| 12 | L301C | G106R | -3.2 | -3.2 | | C+1-/C+5- | 1.7 | 1.7 | 60 | 60 |
| 13 | L302C | G106R | -3.0 | -3.0 | | C+1-/C+5- | 1.9 | 1.9 | 60 | 60 |
| 14 | L302C | G106R | -4.5 | -4.5 | | C+1-/C+5- | 1.8 | 1.8 | 60 | 60 |
| 15 | L302C | G106R | -3.2 | -3.1 | | C+1-/C+5- | 2.3 | 2.5 | 60 | 60 |
| 16 | L302C | G106R | -3.2 | -3.2 | | C+1-/C+5- | 2.0 | 1.8 | 60 | 60 |
| 17 | L302C | G106R | -3.5 | -3.3 | | C+1-/C+5- | 1.8 | 1.8 | 60 | 60 |

**Supplementary Table 3 | DBS electrode information and stimulation parameters for the STN-DBS group.** For “Implantation Depth”, the coordinate origin was defined at the midpoint of the anterior commissure–posterior commissure (AC–PC) line; depths above this reference plan were assigned positive values, whereas depths below were assigned negative values.

| **Number** | **DBS electrodes** | **Neurostimulator** | **Implantation Depth (mm)** | | | **Anode/Cathode** | | **Amplitude (mA)** | | **Pulse width (μs)** | |
| --- | --- | --- | --- | --- | --- | --- | --- | --- | --- | --- | --- |
|  |  |  | **Right** | **Left** | |  |  | **Right** | **Left** | **Right** | **Left** |
| 1 | L301C | G106R | -2.0 | -2.0 | C+1-/C+5- | | 2.5 | | 2.5 | 60 | 60 |
| 2 | L301C | G106R | -1.1 | -1.0 | C+1-/C+5- | | 1.6 | | 1.6 | 60 | 60 |
| 3 | L301C | G106R | -2.0 | -2.0 | C+1-/C+5- | | 2.0 | | 2.0 | 60 | 60 |
| 4 | L301C | G106R | -1.2 | -1.5 | C+1-/C+5- | | 1.5 | | 1.5 | 60 | 60 |
| 5 | L301C | G106R | -1.5 | -1.5 | C+1-/C+5- | | 3.0 | | 3.0 | 60 | 60 |
| 6 | L301C | G106R | -0.9 | -1.3 | C+1-/C+5- | | 3.2 | | 3.2 | 60 | 60 |
| 7 | L301C | G106R | -1.5 | -2.0 | C+1-/C+5- | | 1.7 | | 1.7 | 60 | 60 |
| 8 | L301C | G106R | -2.2 | -2.1 | C+1-/C+5- | | 1.7 | | 1.7 | 60 | 60 |
| 9 | L301C | G106R | -1.5 | -1.5 | C+1-/C+5- | | 2.0 | | 2.0 | 60 | 60 |
| 10 | L301C | G106R | -1.5 | -1.7 | C+1-/C+5- | | 1.7 | | 1.5 | 60 | 60 |
| 11 | L301C | G106R | -2.5 | -1.9 | C+1-/C+5- | | 0.6 | | 0.6 | 60 | 60 |
| 12 | L301C | G106R | -2.0 | -2.4 | C+1-/C+5- | | 1.3 | | 1.3 | 60 | 60 |
| 13 | L302C | G106R | -1.5 | -1.4 | C+1-/C+5- | | 1.2 | | 1.2 | 60 | 60 |
| 14 | L301C | G106R | -1.0 | -1.0 | C+1-/C+5- | | 1.5 | | 1.5 | 60 | 60 |
| 15 | L301C | G106R | -1.9 | -1.5 | C+1-/C+5- | | 1.3 | | 1.5 | 60 | 60 |
| 16 | B33005 | B35300 Percept RC | -1.7 | -1.6 | 2+1-/6+5- | | 2.2 | | 2.2 | 60 | 60 |

**Supplementary Table 4 | Baseline recognition-memory performance in HCs and patients with PD under DBS stimulation inactive.** Recognition-memory performance is summarized for healthy controls and patients with Parkinson’s disease tested under OFF stimulation. Values are presented as mean ± SD. PD-OFF includes all retained PD patients under OFF stimulation, whereas SN-DBS OFF and STN-DBS OFF indicate the two DBS target subgroups under OFF stimulation. HC vs PD-OFF comparisons were used to assess baseline recognition-memory impairment in PD patients relative to healthy controls. SN-DBS OFF vs STN-DBS OFF comparisons were used to assess baseline comparability between the two DBS target groups before active stimulation. q-values indicate Benjamini–Hochberg false-discovery-rate corrected p-values from Welch’s two-sample tests without covariate adjustment.

| **Behavioral measure** | **HC** | **PD-OFF** | **SN-DBS OFF** | **STN-DBS OFF** | **q-value** | |
| --- | --- | --- | --- | --- | --- | --- |
|  |  |  |  |  | **HC vs PD-OFF** | **SN-DBS OFF vs STN-DBS OFF** |
| **d’** | 2.98 ± 0.87 | 1.72 ± 1.20 | 1.55 ± 1.16 | 1.93 ± 1.26 | <0.001 | 0.621 |
| **Pr** | 0.80 ± 0.15 | 0.49 ± 0.32 | 0.44 ± 0.33 | 0.56 ± 0.31 | <0.001 | 0.621 |
| **Hit rate (%)** | 86.2 ± 12.4 | 74.0 ± 20.9 | 74.0 ± 19.1 | 73.9 ± 23.6 | 0.025 | 0.982 |
| **False alarm rate (%)** | 5.8 ± 6.3 | 24.8 ± 22.3 | 30.5 ± 25.4 | 17.9 ± 16.2 | <0.001 | 0.621 |
| **Bias c** | -0.20 ± 0.35 | -0.06 ± 0.58 | 0.03 ± 0.65 | -0.16 ± 0.48 | 0.309 | 0.621 |
| **Hit RT (ms)** | 854 ± 100 | 937 ± 192 | 935 ± 144 | 940 ± 245 | 0.066 | 0.982 |
| **Correct rejection RT (ms)** | 911 ± 116 | 1029 ± 151 | 1016 ± 128 | 1045 ± 178 | 0.010 | 0.826 |

**Supplementary Table 5 | Recognition-memory performance across DBS conditions in patients with PD.** Raw recognition-memory performance is shown separately for the SN-DBS and STN-DBS groups under OFF, 10 Hz and 130 Hz stimulation. Values are presented as mean ± SD. The OFF, 10 Hz and 130 Hz columns show unadjusted descriptive statistics, whereas q values are derived from UPDRS-III-adjusted linear mixed-effects models. The omnibus condition effect tests whether performance differed across OFF, 10 Hz and 130 Hz within each DBS target group. Pairwise contrasts compare stimulation conditions within each DBS target group. qFDR values indicate Benjamini–Hochberg false-discovery-rate corrected p values.

| **Behavioral measure** | **OFF** | **10Hz** | **130Hz** | **Overall stimulation**  **effect, q** | **q-value** | | |
| --- | --- | --- | --- | --- | --- | --- | --- |
|  |  |  |  |  | **OFF vs 10Hz** | **OFF vs 130Hz** | **10Hz vs 130Hz** |
| SN-DBS group | | | | | | | |
| **d’** | 1.55 ± 1.16 | 2.05 ± 0.90 | 2.40 ± 1.09 | 0.005 | 0.011 | 0.002 | 0.429 |
| **Pr** | 0.44 ± 0.33 | 0.62 ± 0.22 | 0.66 ± 0.26 | 0.005 | 0.001 | 0.002 | 0.997 |
| **Hit rate (%)** | 74.0 ± 19.1 | 81.0 ± 13.0 | 80.8 ± 18.1 | 0.177 | 0.195 | 0.311 | 0.509 |
| **False alarm rate (%)** | 30.5 ± 25.4 | 18.7 ± 16.1 | 14.3 ± 14.7 | 0.028 | 0.018 | 0.014 | 0.710 |
| **Bias c** | 0.03 ± 0.65 | -0.04 ± 0.40 | -0.12 ± 0.53 | 0.736 | 0.846 | 0.846 | 0.832 |
| **Hit RT (ms)** | 935 ± 144 | 966 ± 119 | 972 ± 139 | 0.736 | 0.880 | 0.880 | 0.964 |
| **Correct rejection RT (ms)** | 1016 ± 128 | 1081 ± 157 | 1065 ± 143 | 0.479 | 0.639 | 0.793 | 0.793 |
| STN-DBS group | | | | | | | |
| **d’** | 1.93 ± 1.26 | 1.40 ± 1.10 | 1.65 ± 1.35 | 0.172 | 0.036 | 0.236 | 0.480 |
| **Pr** | 0.56 ± 0.31 | 0.42 ± 0.31 | 0.46 ± 0.36 | 0.172 | 0.035 | 0.075 | 0.997 |
| **Hit rate (%)** | 73.9 ± 23.6 | 72.9 ± 23.1 | 71.7 ± 20.1 | 0.669 | 0.497 | 0.311 | 0.509 |
| **False alarm rate (%)** | 17.9 ± 16.2 | 31.2 ± 26.6 | 25.2 ± 24.2 | 0.172 | 0.042 | 0.201 | 0.642 |
| **Bias c** | -0.16 ± 0.48 | 0.09 ± 0.69 | -0.10 ± 0.52 | 0.451 | 0.846 | 0.846 | 0.754 |
| **Hit RT (ms)** | 940 ± 245 | 979 ± 251 | 976 ± 207 | 0.590 | 0.880 | 0.880 | 0.964 |
| **Correct rejection RT (ms)** | 1045 ± 178 | 1029 ± 177 | 1071 ± 233 | 0.803 | 0.793 | 0.855 | 0.846 |

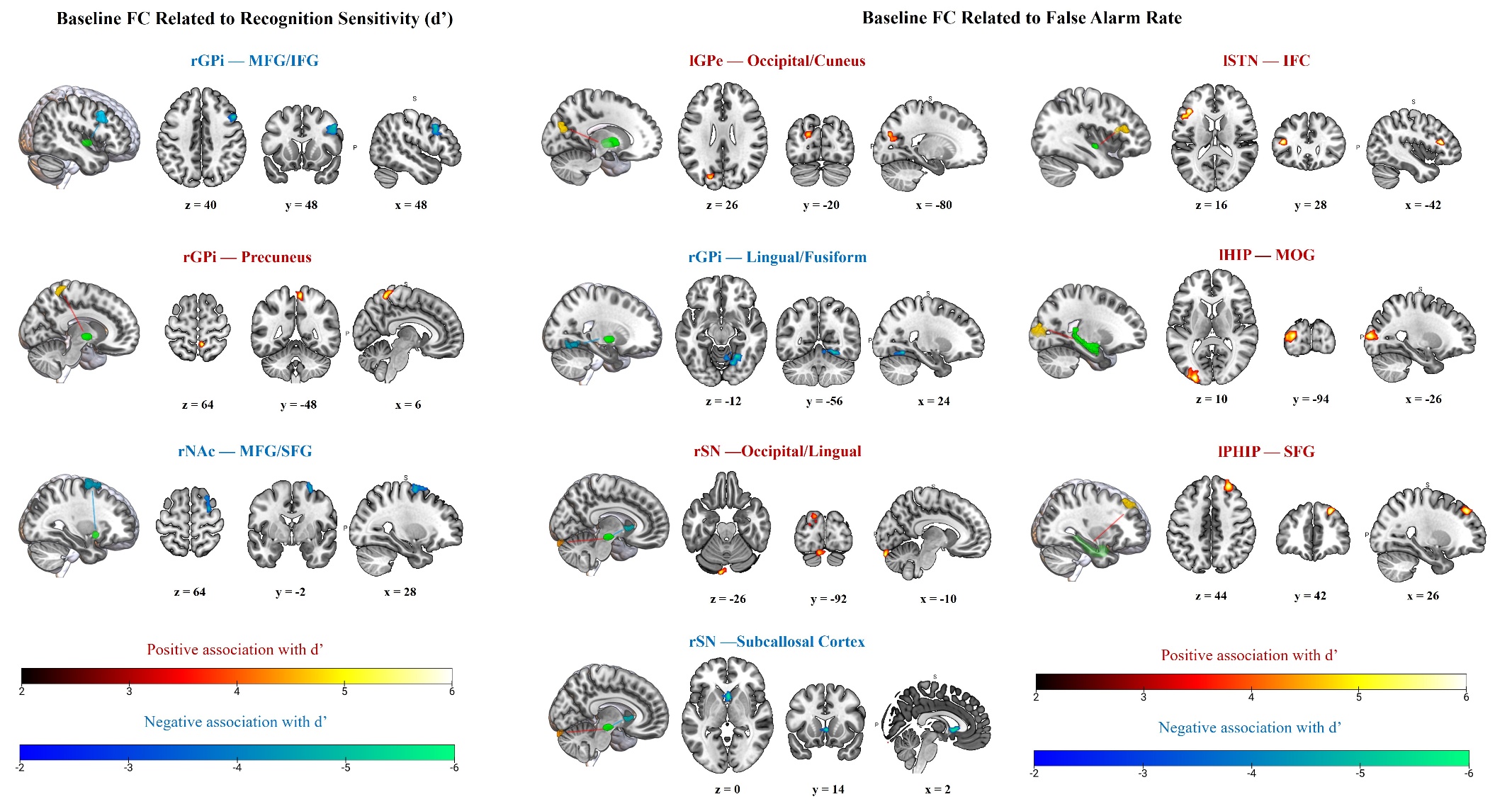

**Supplementary Figure 3 | Baseline resting-state functional connectivity associated with recognition performance under DBS-OFF.** Baseline seed-to-voxel resting-state functional connectivity analyses were performed under the DBS-OFF condition to identify intrinsic connectivity patterns associated with recognition-memory performance. Recognition sensitivity, indexed by d′, was associated with pallidal and accumbens connectivity with frontal and posterior cortical regions. Lower d′ was related to stronger right GPi connectivity with right middle/inferior frontal cortex and stronger right nucleus accumbens connectivity with right middle/superior frontal cortex, whereas higher d′ was related to stronger right GPi connectivity with the precuneus. False alarm rate was associated with a broader false-recognition network involving basal ganglia, nigral, medial temporal, frontal and posterior visual regions. Higher false alarm rate was associated with stronger left GPe–occipital/cuneus, right SN–occipital/lingual, left STN–inferior frontal cortex, left hippocampus–middle occipital gyrus and left parahippocampal gyrus–superior frontal gyrus connectivity. Lower false alarm rate was associated with stronger right GPi–lingual/fusiform and right SN–subcallosal cortex connectivity. Warm colors indicate positive associations with behavioral measures, whereas cool colors indicate negative associations. Coordinates indicate MNI slice locations. Imaging results were thresholded at voxel-wise p < 0.001 with cluster-level FWE correction at p < 0.05.

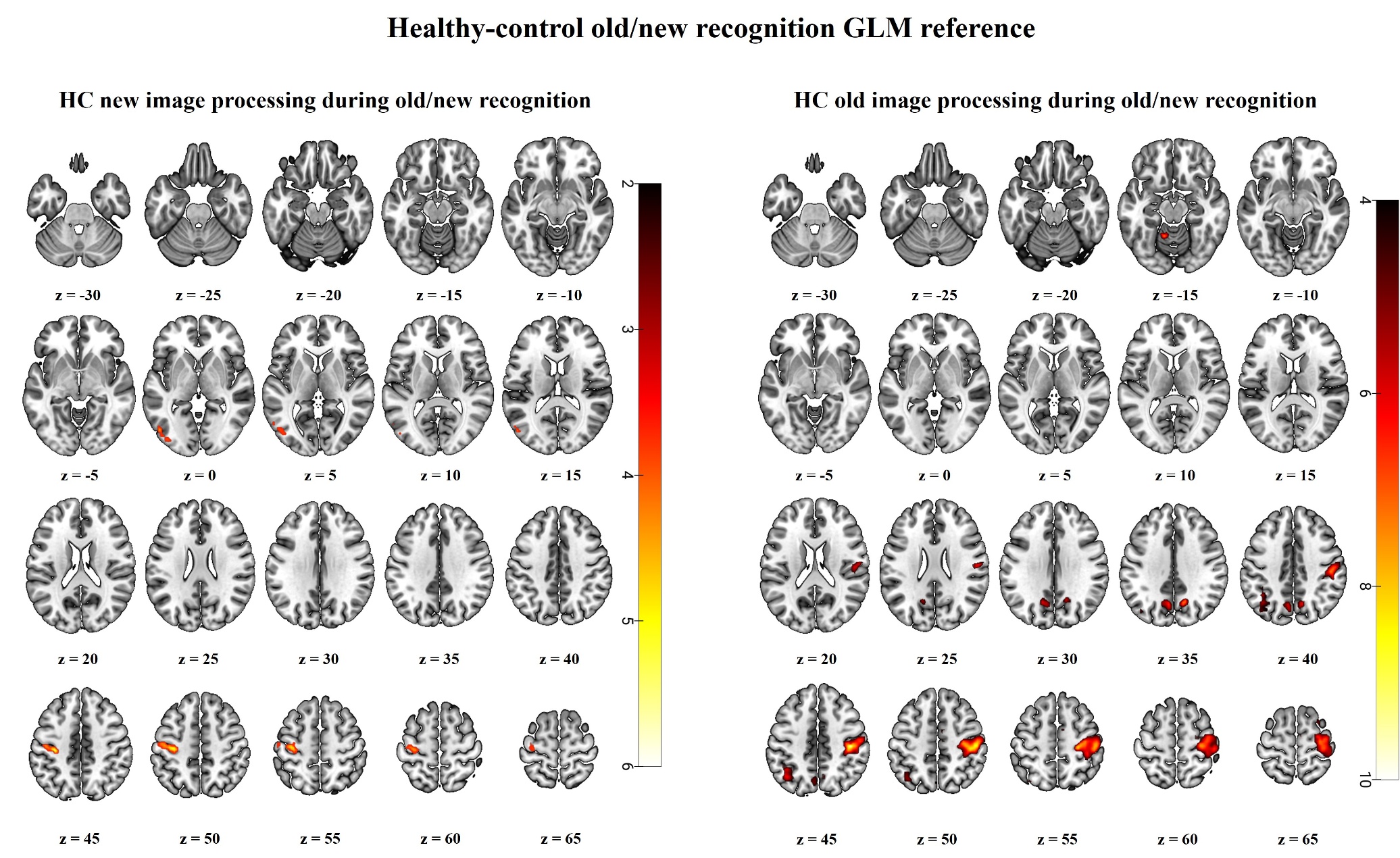

**Supplementary Figure 4 | Healthy-control new-image processing during old/new recognition.** Left panel: HC new image processing during old/new recognition. Whole-brain one-sample GLM result in healthy controls showing greater activation for valid new-image retrieval trials than valid old-image retrieval trials. The contrast was defined as New > Old. Significant clusters were observed in left lateral occipital/occipito-temporal cortex and left precentral/postcentral cortex. Right panel: HC old image retrieval during old/new recognition. Whole-brain one-sample GLM result in healthy controls showing greater activation for valid old-image retrieval trials than valid new-image retrieval trials. The contrast was defined as Old > New. Significant clusters involved bilateral precuneus/cuneus, left inferior parietal/angular cortex, right precentral/postcentral cortex, supplementary motor area and superior frontal/premotor regions. Images are thresholded at voxel-wise p < 0.001 uncorrected with cluster-level FWE correction at p < 0.05.

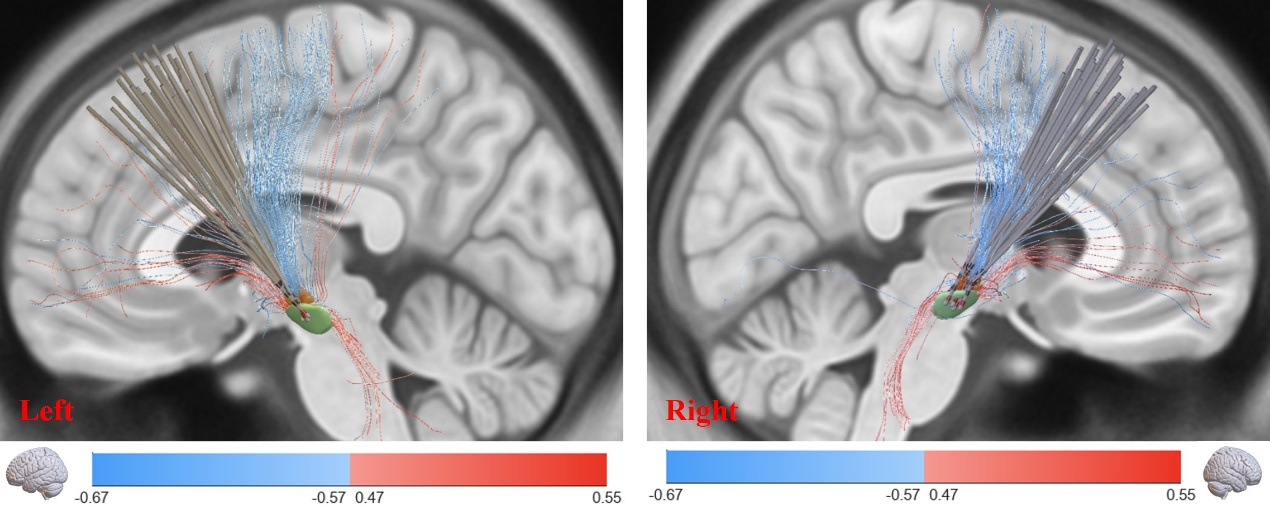

**Supplementary Figure 5 | Connectome-based fiber filtering of 10 Hz–related changes in overall recognition accuracy.** Streamlines whose engagement covaried positively (red) or negatively (blue) with the change in overall recognition accuracy at 10 Hz relative to OFF-state were identified using Lead-DBS fiber filtering on the PPMI PD connectome (ppmi_85_ewert_2017) with mirrored hemispheres. Positively weighted streamlines projected predominantly toward prefrontal regions, whereas negatively weighted streamlines targeted sensorimotor cortex, including the postcentral gyrus with extension into the precentral gyrus. Using preoperative T1-weighted MRI and postoperative CT, we reconstructed DBS lead locations and estimated stimulation fields in standardized template space. To relate stimulation anatomy to cognitive outcomes under low-frequency stimulation, we quantified each patient’s change in overall recognition accuracy at 10 Hz relative to OFF-state and performed connectome-based fiber filtering in Lead-DBS v3.024 (https://www.lead-dbs.org), a MATLAB-based toolbox (R2022b, MathWorks, Natick, MA, USA), using the PPMI patient connectome (PPMI_85_ewert_2017), with hemispheres mirrored to reduce lateralization-related variance. This workflow identifies streamlines consistently engaged by patients’ VTAs and assigns each streamline a selectivity statistic (“Fiber T-score”) by comparing behavioral change between VTAs connected versus not connected to that streamline. At an exploratory, uncorrected threshold (α < 0.05), we identified 54 positively weighted streamlines and 207 negatively weighted streamlines. Positively weighted streamlines predominantly projected toward prefrontal regions, whereas negatively weighted streamlines preferentially projected toward sensorimotor cortices, including the postcentral gyrus and the precentral gyrus, with a prominent parietal component. These findings suggest that engagement of dissociable cortico-subcortical ganglia pathways accompany cognitive improvement versus deterioration under 10 Hz stimulation. To validate robustness, we performed permutation testing of the association between model-based estimates and empirical behavioral change, which remained highly significant (Spearman R = 0.71, P_perm = 2.0 × 10^−4; Pearson R = 0.70, P_perm = 4.0 × 10^−4; [N_perm = 1000]).
